# Association of genetic variation at the GJA5/ACP6 locus with motor progression in Parkinson’s

**DOI:** 10.1101/2022.10.28.22281645

**Authors:** Alejandro Martinez-Carrasco, Raquel Real, Michael Lawton, Regina H Reynolds, Manuela M. X. Tan, Lesley Wu, Nigel M. Williams, Camille Carroll, Jean-Christophe Corvol, Michele T.M. Hu, Donald G. Grosset, John Hardy, Mina Ryten, Yoav Ben-Shlomo, Maryam Shoai, Huw R. Morris

## Abstract

**Importance:** There is a pressing need to understand the biology of Parkinson’s disease (PD) progression and to identify biological pathways as possible therapeutic targets.

**Objective:** To identify genomic variation associated with PD motor presentation and early stage progression.

**Design:** GWAS meta-analysis of early PD motor progression, from multiple longitudinal cohorts, using MDS-UPDRS III clinical assessments.

**Setting:** Multicentre

**Participants:** 3572 unrelated European ancestry individuals diagnosed with PD from 6 studies.

**Main Outcomes and Measures:** Linear mixed effect models under an additive model corrected for age at diagnosis, gender, and the first 5 genetic principal components (PCs), with axial, limb, and total MDS-UPDRS III as outcomes of the model.

**Results:** We identified an association between the PD axial rate of progression and variation at the *GJA5* locus at 1q12 (Beta = -0.25, SE = 0.04, *P* = 3.4e-10). Exploration of the regulation of gene expression in the region (cis-eQTL analysis) showed that the lead variant was associated with expression of *ACP6*, a lysophosphatidic acid phosphatase that regulates mitochondrial lipid biosynthesis (cis-eQTLs p-values in blood and brain RNA expression datasets: < 10^−8^ in eQTLGen and 10^−7^ in PsychEncode). In addition, we found a nominal association between axial motor presentation and the *MAD1L1* gene at 7q21.11 (Beta = 0.54, SE = 0.11, *P* = 1.6e-7), a gene previously associated with neuropsychiatric disease, and in the long non-coding RNA *LINC00511* at 17q21.31 (Beta = -0.62, SE = 0.11, *P* = 6.3e-8). Further functional annotation allowed us to nominate the likely causal variants and determine that variants at 7q21.11 may be related to dysregulation of *MAD1L1* expression. Variants at *LINC00511* may cause a disruption of a distal epigenetic regulation of *SOX9* through an anchored chromatin loop.

**Conclusions and Relevance:** Our large multicentre study sheds new light on the genetic architecture of PD progression, which is distinct from PD susceptibility. Our study highlights the potential role of mitochondrial lipid homeostasis in the progression of PD, which may be important in establishing new drug targets to tackle disease progression.

## Introduction

Parkinson’s disease (PD) is a progressive neurodegenerative disorder with motor and non-motor symptoms, clinically manifesting with rigidity, postural instability, and slowness of movement (bradykinesia). The motor deficits are linked to the loss of dopaminergic (DA) neurons in the substantia nigra pars compacta, their projection to the striatum, and the accumulation of alpha-synuclein aggregates in Lewy bodies in the remaining basal ganglia neurons ^1^.

PD is heterogeneous in its progression and onset. The predominant motor phenotype is influenced by age at PD onset, with tremor being more prominent in advanced age, and the risk of dystonia at presentation increasing in younger individuals ^2^. With respect to progression, younger patients tend to have a slower rate of motor progression as measured using the MDS-UPDRS and Hoehn and Yahr assessments ^3^. Functional imaging studies have also shown a slower rate of decline in the loss of nigrostriatal terminals in early onset compared to late onset PD ^4^.

To date, the majority of PD genetic studies have focused on the risk of developing PD^5^. Relatively little is known about the genetic factors that contribute to variation in the onset and progression of motor and non-motor symptoms. Studies of PD age at onset have shown that the genetic determinants of age at onset are different to the genetic factors determining case-control status, with the *MAPT* and *GCH-1* loci increasing disease risk but not associated with age at onset ^6–8^. With respect to common variability explaining differences in the disease progression, Iwaki and colleagues performed a large GWAS looking at 25 different outcome measures in a meta-analysis of 12 longitudinal cohorts, including mortality, dementia, disease severity, and patient disability^9^.

They reported an association between an intronic variant in *SLC44A1*, a mitochondrial choline transporter, and motor progression, reflected by reaching a Hoehn and Yahr score higher or equal than 3 (HY3). A more recent study carried out by Tan and colleagues took a different approach using a principal components (PC) based measure that combined multiple assessments for composite motor and cognitive progression. They found a novel association between *ATP8B2* and PC based motor progression in a gene-based analysis ^10^.

These studies suggest that pathways specifically related to the progression of PD can be understood from genotype/phenotype analysis, which may ultimately lead to the development of new disease modifying therapies. We modelled the early stages of motor Parkinson’s disease, using the total score from the MDS-UPDRS part III, a validated scale recommended for clinical trials to measure both response to levodopa treatment and the rate of change over time ^11^. We have also derived and analysed axial and limb motor stages from the scale, as they may relate in part to separate pathology ^12^. We have used a genome wide association (GWA) approach to define genetic determinants associated with variation in motor severity and those causing differences in motor progression. Finally, we have performed functional annotation and fine-mapping to understand how the nomicated genetic variants are associated with the underlying biology of PD motor traits.

## Key points

### Question

Do genetic variants explain part of the high variability in the rate of change of clinical motor assessments in Parkinson’s disease across observational studies and clinical trials?

### Findings

This genome-wide association meta-analysis identified genetic variability explaining differences in the progression of the MDS-UPDRS axial motor score. Variation at the 1q12 locus, which determines the level of expression of the ACP6 gene, was significantly associated with a lower axial rate of change and this finding was replicated in each cohort included in this large genetic study.

### Meaning

We have identified evidence that variation in mitochondrial lipid biosynthesis may relate to PD progression and this may help to define new treatments that modify the disease course and in the design of future studies

## Methods

### Study design and quality control at the sample and genetic level

We studied six PD observational and interventional longitudinal cohorts with either genotyping or whole genome sequencing (WGS) data available, totalling 4971 patients (eTable 1 in Supplement).

We selected cohorts which included longitudinal MDS-UPDRS part III examination from the Movement Disorder Society - Unified Parkinson’s disease Rating Scale (MDS-UPDRS)^11^, and applied quality control (QC) at the clinical data level (eFigure 1 in Supplement).

To study the motor progression of PD we derived limb and axial phenotypes from the MDS-UPDRS part III scale based on previously accepted definitions^13^. In addition, we used the MDS-UPDRS III total score as an overall measure of PD motor state (eTable 2 in Supplement). We included data up to 36 months from the baseline visit, within longitudinal observational and therapeutic cohorts, as we had high rates of data completion up to 36 months.. Imputation of patients’ missing motor outcomes was performed when possible (eMethods in Supplement; eTable 3 in Supplement).

We applied genetic QC at the sample and variant level followed by imputation in the Michigan Imputation Server (RRID:SCR_017579; https://imputationserver.sph.umich.edu) ^14^, and post-imputation QC (eMethods ; eFigure 1 in Supplement). Table 1 summarises the demographics of the data after QC.

**Table 1.** Cohort demographics and motor scores percent rate of change.

| Study | N patients<sup>a</sup> | N Observations<sup>b</sup> | Visit interval, mo | ON/OFF | No. (%) male | AAD, years mean(sd) | AAB, years mean(sd) | Duration<sup>c</sup>, years mean(sd) | Total yearly rate of change mean (sd) | Limb yearly rate of change mean (sd) | Axial yearly rate of change mean (sd) |
| --- | --- | --- | --- | --- | --- | --- | --- | --- | --- | --- | --- |
| TPD | 1699 | 4349 | 18 | ON | 1101 (64.8) | 66.20 (±9.24) | 67.50 (±9.31) | 3.12 (±3.04) | 2.7 (±4.69) | 1.81 (±3.2) | 0.88 (±1.35) |
| OPDC | 797 | 1978 | 18 | ON | 513 (64.37) | 66.04 (±9.46) | 67.25 (±9.57) | 2.86 (±1.84) | 2.85 (±4.27) | 1.9 (±3.56) | 0.95 (±1.29) |
| PPMI | 287 | 1653 | 3 & 6 & 12 | OFF | 184 (64.11) | 61.01 (±9.73) | 61.59 (±9.70) | 2.02 (±1.97) | 3.01 (±3.65) | 2.36 (±2.98) | 0.65 (±1.10) |
| PD STAT | 124 | 358 | 12 | OFF | 76 (61.29) | NA | 66.08 (±9.37) | 10.14 (±4.06) | 1.70 (±5.76) | 0.88 (±4.83) | 0.80 (±1.90) |
| DIGPD | 305 | 1005 | 12 | ON | 184 (60.33) | 59.68 (±9.85) | 62.59 (±9.70) | 3.74 (±1.55) | 2.37 (±3.41) | 1.81 (±2.91) | 0.53 (±0.95) |
| PDBP | 360 | 2090 | 6 | ON | 222 (61.67) | 59.9 (±10.90) | 64.73 (±9.15) | NA | 1.05 (±4.11) | 0.63 (±3.47) | 0.41 (±1.32) |

Abbreviations = N, Number; AAD, Age at Diagnosis; AAB, Age at Baseline,
<sup>a</sup>N patients = Number of PD patients after QC was applied
<sup>b</sup>N observations = Number of observations including MDS-UPDRS part III motor assessment after QC was applied
<sup>c</sup>Duration = Disease duration from onset to study entry

Previous studies have reported that levodopa improves motor state examination and may slow disease progression^15^. Because the motor improvement is noticeable a few hours after treatment, having an impact on the MDS-UPDRS measures, we have compared individuals from cohorts in the same state at each assessment. If cohorts had data in the “OFF” state available, we used longitudinal “OFF” vs “OFF” MDS-UPDRS part III scores, otherwise “ON” vs “ON”. In addition, we performed a sensitivity analysis adjusting the motor states by levodopa usage (eMethod in supplement).

### Statistical approaches

To study the effect of genetics on motor progression as well as on baseline variability, we used Linear Mixed Effect Models (LMMs) which can be used to study longitudinal measures as the variability in the outcome can be studied at two levels, within groups (i.e. individual patient visits), or between groups (across patients).

After performing a thorough model comparison based on different combinations of predictors, we nominated two models (A and B). In model A, we studied the additive genetic effect of SNPs on the average motor outcome. With model B we studied the additive effect SNPs have on the outcomes’ rate of change (eMethods in Supplement). Both models A and B met the LMMs model assumptions (eFigure 3 in Supplement). We quantified the power of using LMMs to investigate the effect of SNPs in changes in the motor signs of PD (eMethods in Supplement).

To run model A, we used lmerTest R package (v. 3.1-3; RRID:SCR_015656 ; https://cran.r-project.org/web/packages/lmerTest/lmerTest.pdf) and the Satterthwaite approach to approximate degrees of freedom and derive p-values, and fitted them using restricted maximum likelihood (REML), as it produces acceptable Type-1 error rates^16^. In addition, we used SCEBE^17^ algorithm (v. 0.1.0; https://github.com/Myuan2019/SCEBE) with REML and lme4 R package (v. 1.1-30; RRID:SCR_015654; https://cran.r-project.org/web/packages/lme4/index.html) to decrease the computational expense of adding unexplained variability at the slope level in model B (eMethods in Supplement). We validated SCEBE in two separate cohorts (eFigure 4 in Supplement). All tests were two-tailed. We used METAL software (version released on the 25/03/2011; RRID:SCR_002013; http://csg.sph.umich.edu//abecasis/Metal/) for meta-analysis of genome wide association summary statistics, using a fixed effects model weighted by β coefficients and the inverse of the standard errors^18,19^. We also applied QC to the meta-analysis results (eMethods in Supplement). Statistical significance was assessed at the genome-wide level (P = 5 × 10^−8^).

### Fine Mapping and functional annotation

We performed a conditional and a stepwise model selection procedure to determine if there were independently associated SNPs in each locus of interest. To nominate causal variants, we applied fine-mapping. To determine whether causal mutations could be linked to the motor phenotypes by dysregulation in expression, we performed co-localization against cis-eQTL datasets. To further understand regulatory mechanisms in nominated genomic regions, we mapped each locus to the brain cell type specific enhancer-promoter interactome data generated by Nott and colleagues, and to regulatory elements data from the FANTOM5 (RRID:SCR_002678) project ^20,21^. Software and packages to conduct the analyses are described in eMethods in Supplement. In addition, we used FUMA, a web-based platform that integrates a wide range of functional annotation data (RRID:SCR_017521; version 1.3.8; https://fuma.ctglab.nl/) ^22^.

## Results

The overall change in MDS-UPDRS III total score was similar in the TPD, OPDC, PPMI, and DIGPD cohorts at 2.37-3.01 points/year. PD-STAT and PDBP had a lower rate of change for the total and limb motor scores, which we hypothesised was related either to longer disease duration, or to selection effects related to “benign” PD in patients with longer disease duration. We assessed this by fitting a LMM using data from TPD, and found a significant interaction between time and disease duration in determining MDS-UPDRS progression (Beta = -0.11, SE = 0.04, *P* = 0.01). In this data, longer disease duration was associated with a slower rate of change in MDS-UPDRS, which appears to be non-linear with extended disease durations. The axial rate of change was more consistent across cohorts, except for PDBP which had a lower rate of change for all motor scores (Table 1). Overall, we confirmed that the MDS-UPDRS measures increased, reflecting worsening motor impairment, from study entry up to 3 years (Figure 1).

**Figure 1.**
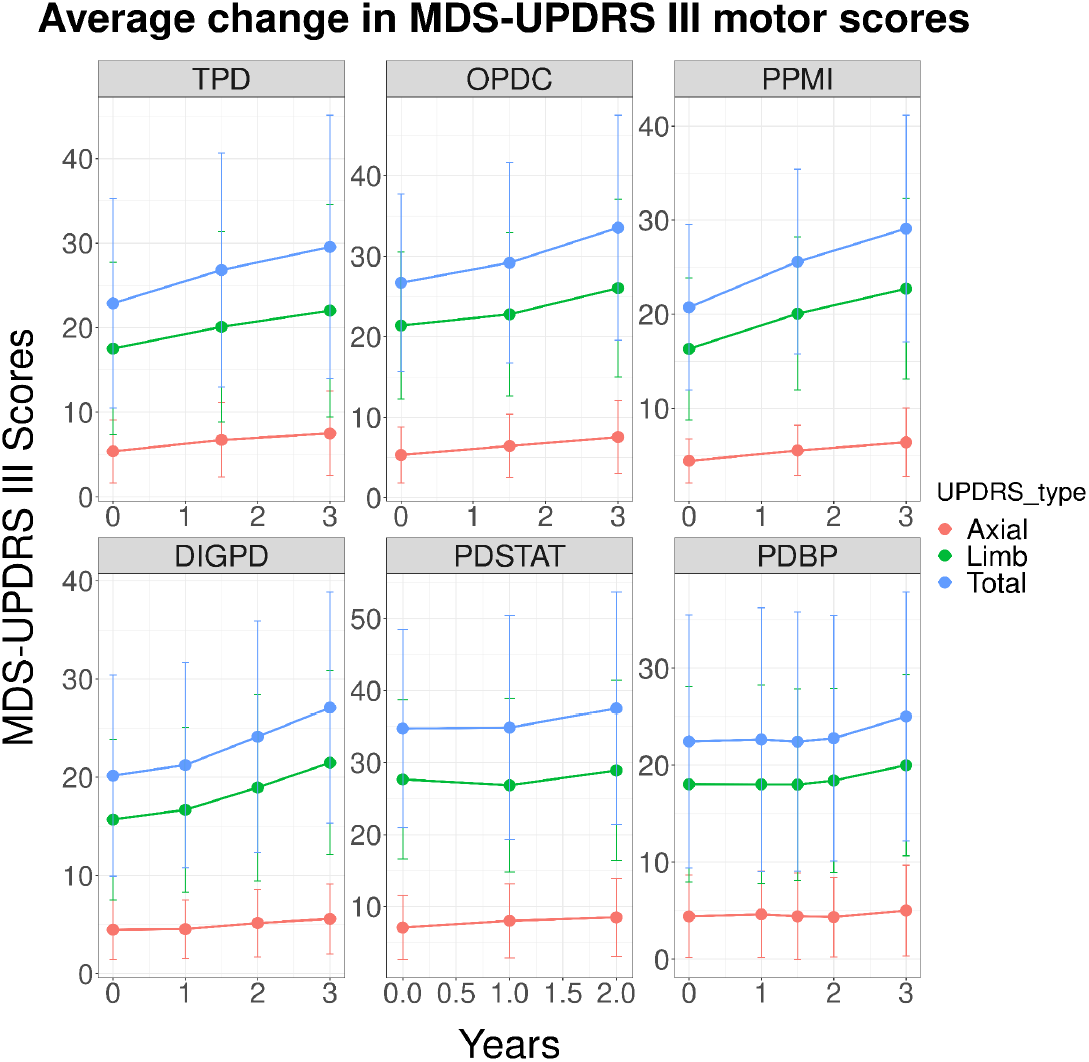
Trajectory of the MDS-UPDRS III motor scores across cohorts. In the X axis, the time point at which the MDS-UPDRS III assessment was measured. Each plot shows the motor scores trajectories on each cohort highlighted in the label. The y axis represents the average scores for each of the motor states. The bars represent the SD of the average motor scores.

Our power calculation showed that the current LMM was well powered to detect high effect sizes (Beta ≥ 0.2) for a wide range of different MAFs, with a limit for variants with an allele frequency ≤ 1% (eFigure 5 in Supplement).

We performed a GWAS with models A and B, and meta-analysed results separately allowing for genomic control to correct the test statistics of those cohorts that had genomic inflation (λ > 1 & λ < 1.2) (eTable 4 in Supplement).

We found two LD blocks approaching genome wide significance associated with the overal axial motor severity (*MAD1L1* in chromosome 7 and *LINC00511* in chromosome 17), and one haplotype block associated with the axial rate of progression (*GJA5* in chromosome 1) (Figure 2a,b). The lead SNP in *MAD1L1* was rs4721411 (Beta = 0.54, SE = 0.11, *P* = 1.6e-7), and the lead variant in the long non-coding RNA *LINC00511* was rs36082764 (Beta = -0.62, SE = 0.11, *P* = 6.3e-8) (eTable 5 in Supplement). Although the lead variant in the *GJA5* locus was not assayed in the PPMI data, the significant proxy variant rs12037169 (Beta = -0.25, SE = 0.04, *P* = 3.93e-10) was present in all cohorts. We found the association test-statistic and directionality of each of these variants to be consistently replicated across cohorts (eTable 6 in Supplement) (Figure 2c-2e).

**Figure 2.** GWAS meta-analysis of motor axial presentation and rate of change. (Figure 2 a,b,c,d and e will be configured as a composite figure if accepted for submission)

**Figure 2a.**
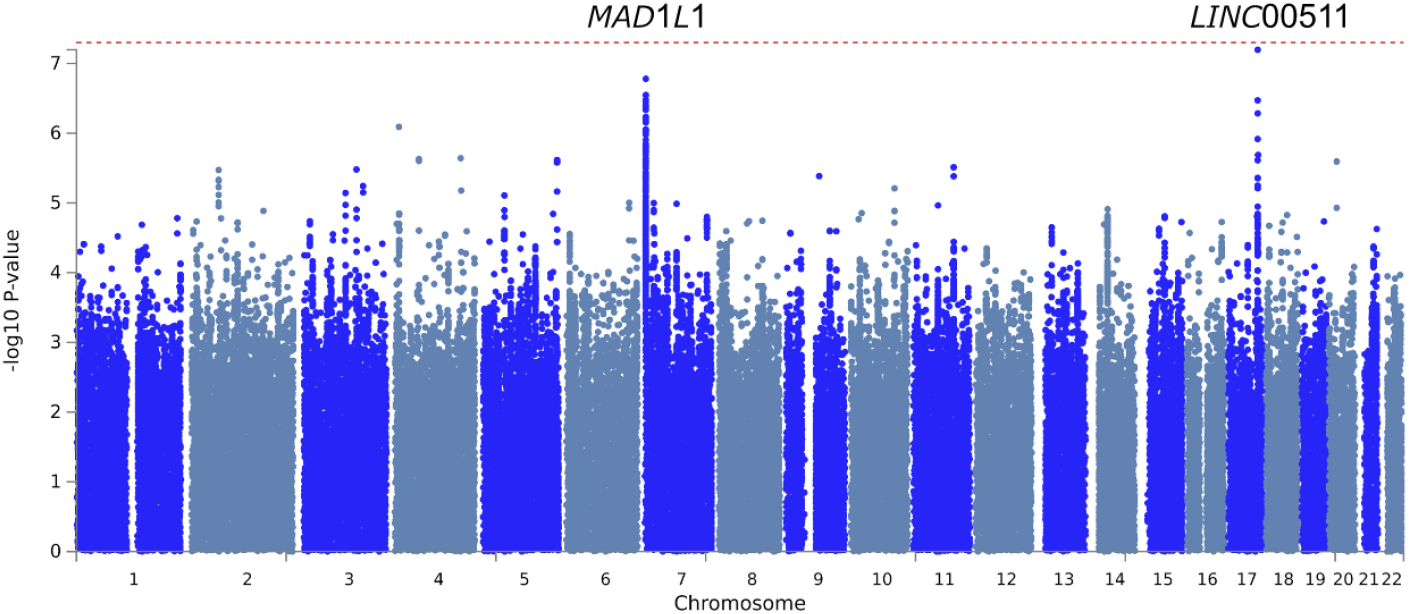
Manhattan plot for the axial severity GWAS meta-analysis. The red dashed line indicates the genome-wide significance threshold P-value = 5e-8. *MAD1L1* is highlighted on chromosome 7 and *LINC00511* on chromosome 17.

**Figure 2b.**
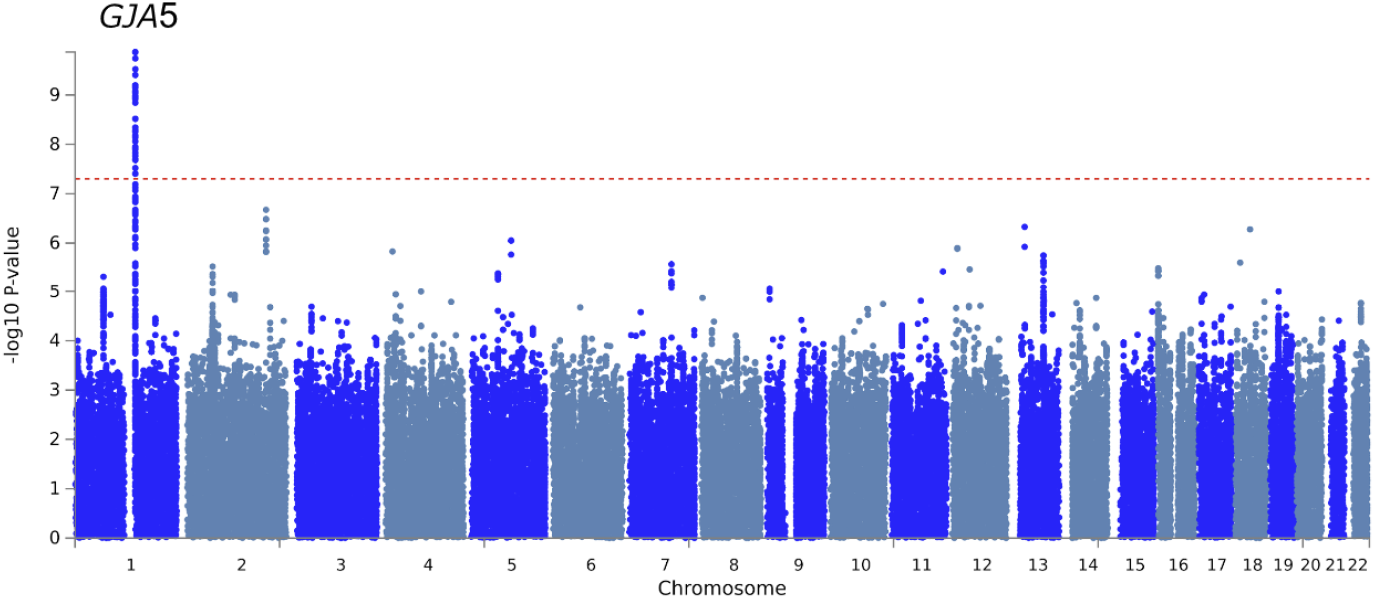
Manhattan plot of the rate of axial change GWAS meta-analysis. On the X axis each of the 22 chromosomes, and each SNP P-value on the Y axis. The red dashed line indicates the genome-wide significance threshold P-value = 5e-8. *GJA5* is highlighted on chromosome 1. Each dot corresponds to the P-Value of the conditional likelihood interaction term between SNP and time (SNP*time).

**Figure 2c.**
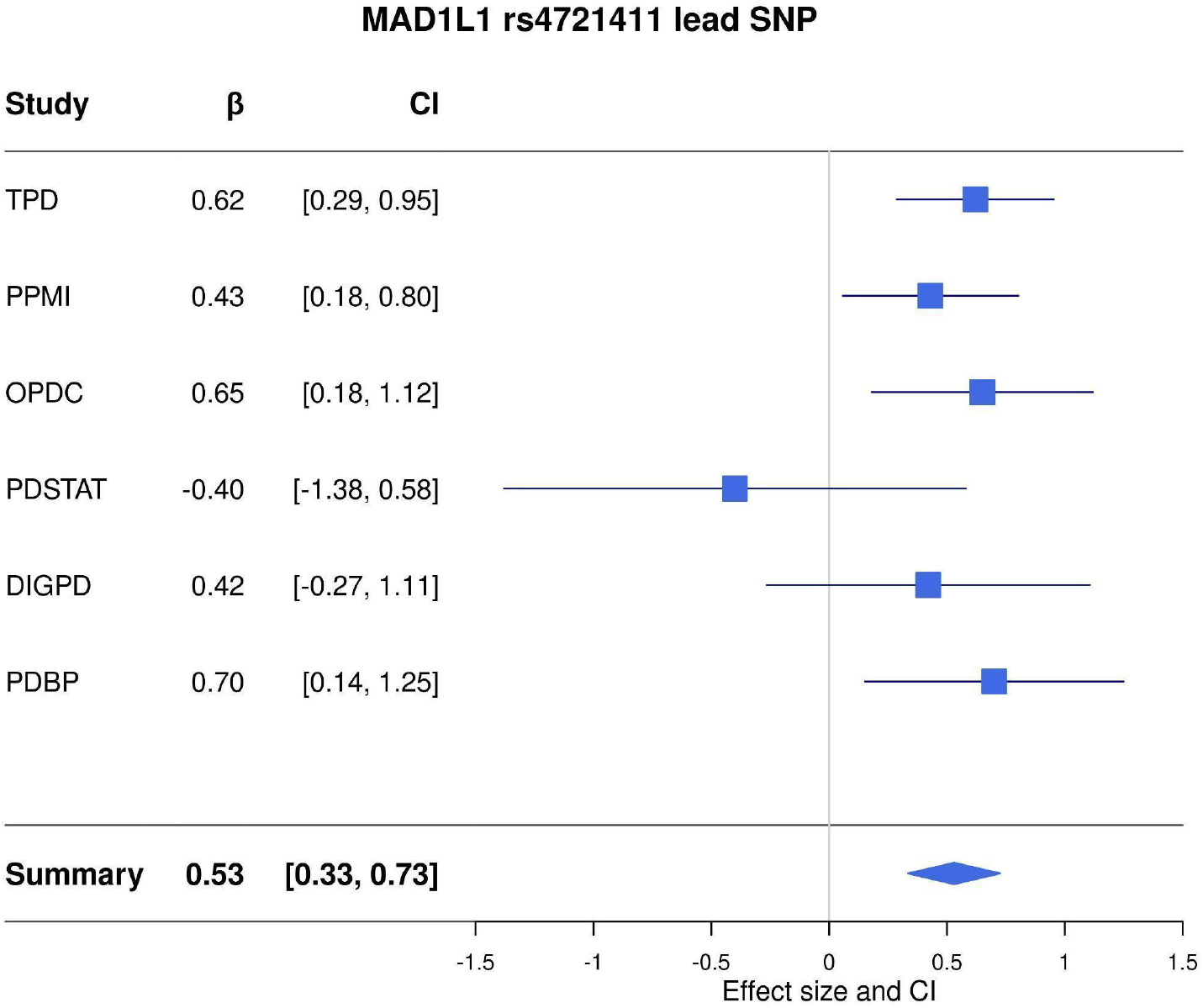
Forest plots for lead variant rs4721411-T found at the MAD1L1 locus (right) under model A (I^2^ = 0; Cochran’s Q test: ꭓ^2^ = 4.01, df = 5, *P* =0.55) annotated by study, effect size, and the corresponding 95% confidence interval.

**Figure 2d.**
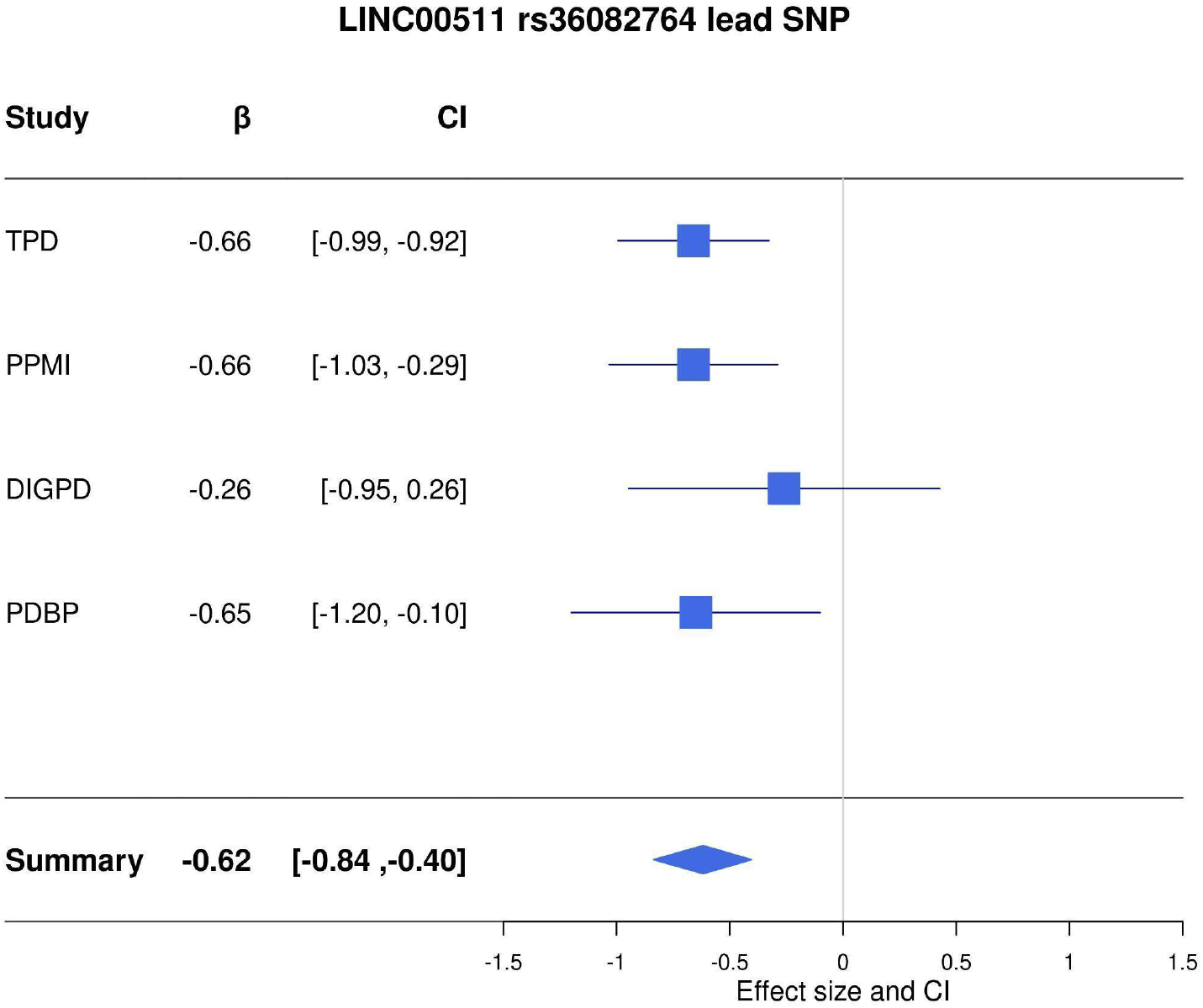
Forest plots for lead variant rs36082764-T found at *LINC00511* locus under the GWAS meta-analysis using model A (I^2^ = 0; Cochran’s Q test: ꭓ^2^ = 1.07, df = 3, *P* = 0.78) annotated by study, effect size, and the corresponding 95% confidence interval.

**Figure 2e.**
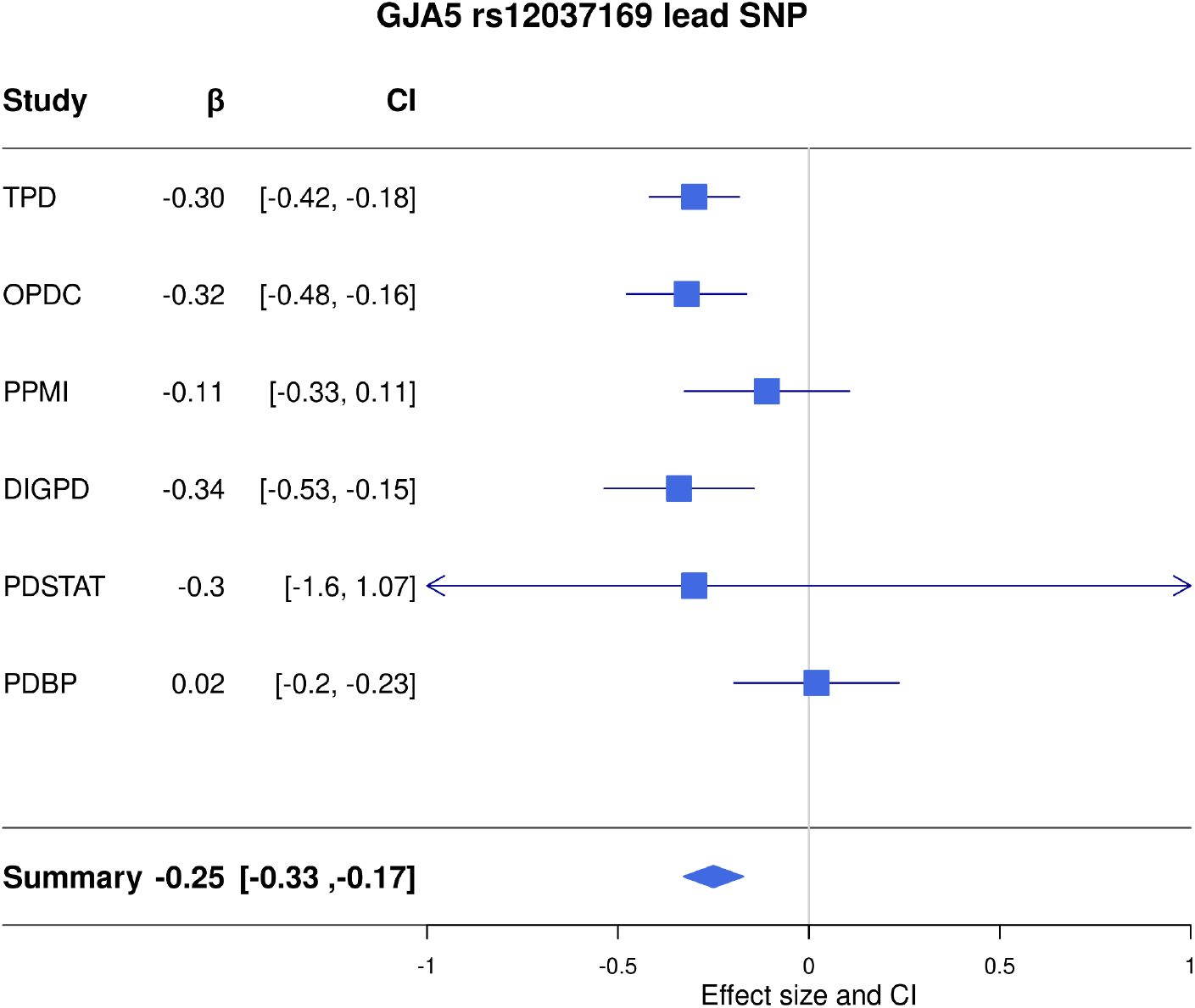
Forest plots for lead variant rs12037169 within GJA5 under the GWAS meta-analysis using model B for the axial outcome (I^2^ = 40.1; Cochran’s Q test: ꭓ^2^ = 9.64, df = 5, *P* = 0.10), annotated by study, effect size, and the corresponding 95% confidence interval.

For rs36082764, we found the effect to be consistent across cohorts, although it was not present in data from PD-STAT and OPDC. For OPDC, we checked all SNPs in LD with this variant, and we found two variants significantly associated in the same direction (rs7216312 Beta = -0.50, SE = 0.25, *P* = 0.05 D’ = 1; rs7218330 Beta = -0.48, SE = 0.25, *P* = 0.048, D’ = 0.99).

We then investigated if there were independently associated SNPs at each locus. We did not find any signal other than the lead SNPs in the selection procedure, using conditional and stepwise selection procedure analyses using GCTA-COJO. Under a single causal variant assumption, we then performed statistical fine mapping. We nominated rs3778978 in *MAD1L1* and rs7213651, rs7218929, and rs12950478 in *LINC00511* as the consensus SNPs as reported across fine-mapping tools. (eFigure 6 in Supplement ; eTable 7 in Supplement). We did not find any consensus SNPs at the *GJA5* locus.

We investigated the potential effects of these GWAS loci in controlling gene expression, using available genomic regulatory elements datasets. We mapped them against: i.) cell type specific and “bulk” enhancer marks, ii.) enhancer-transcription start site interaction marks from FANTOM5, and iii.) brain cell type-specific transcriptional regulatory marks and distal enhancer-promoter interactions. We found that the *MAD1L1* fine-mapped causal variant and the lead SNP overlapped with an active enhancer mark, and this region was predicted to interact with a transcription start site (TSS), supporting an effect of the GWAS nominated variants in regulating the expression of *MAD1L1* (Figure 3a). For *LINC00511*, we found an anchored chromatin loop from the GWAS LD block in *LINC00511* to a region where the *SOX9* neuronal active promoter is found, suggesting that mutations in this distal regulatory region may alter *SOX9* expression in neurons specifically (Figure 3b). We did not find any suggestive regulatory event in the *GJA5* locus based on data from the FANTOM5 and Nott studies.

**Figure 3.** Functional annotation of nominated loci in GWASs meta-analysis. (Figure 3 a,b,c will be configured as a composite figure if accepted for submission)

**Figure 3a.**
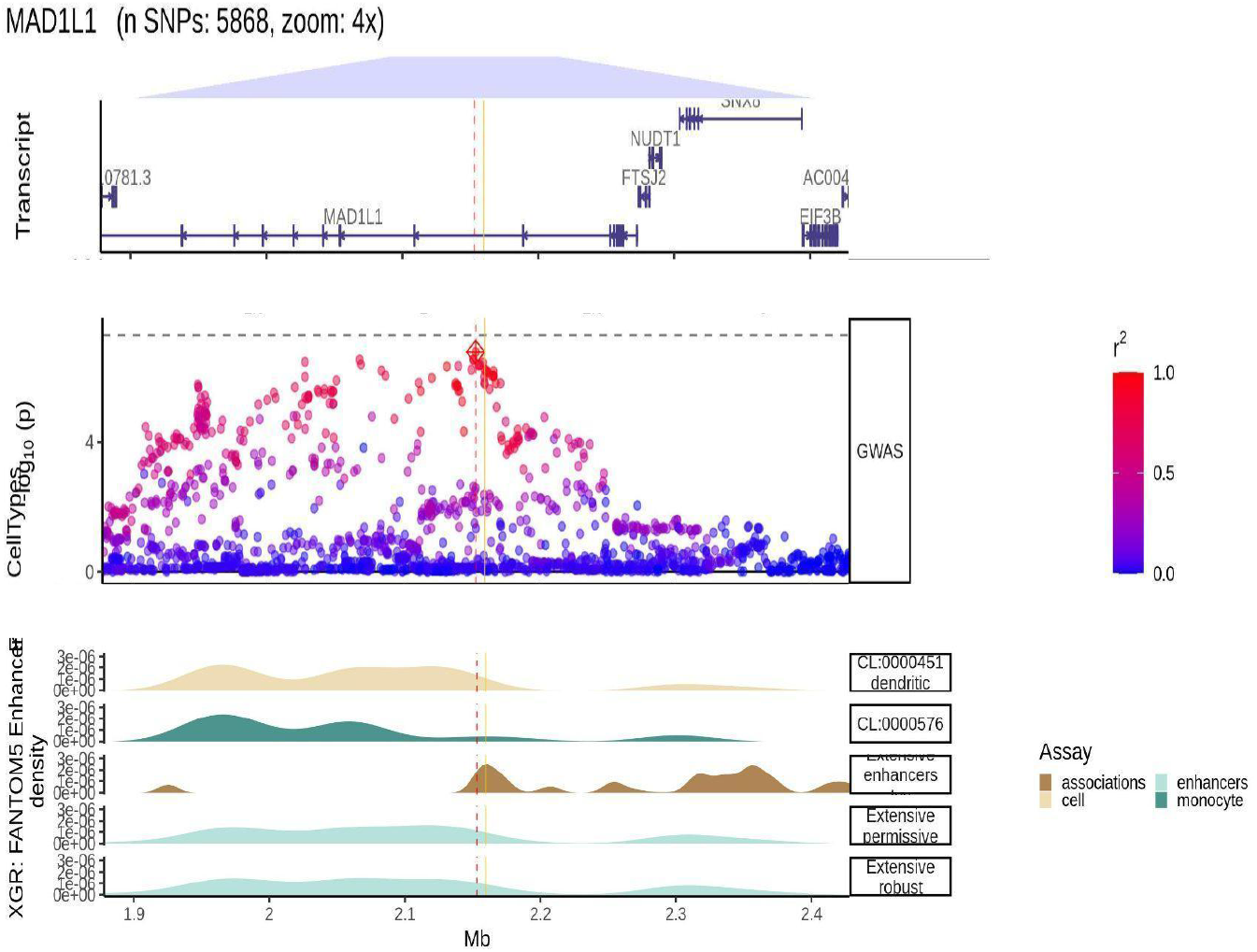
Transcripts plot (top), locus plot (middle), and FANTOM5 enhancer marks from the FANTOM project. All data from the FANTOM5 project was scanned to plot out the 5 datasets with present enhancer marks on the region of interest. From top to bottom, dendritic and monocytes cell type specific enhancer marks, bulk enhancer transcription start site interaction mark, and bulk enhancer permissive and robust marks. In the locus plot from the middle, the SNPs are coloured in red as LD (given by R2) increases, and blue as the LD decreases. From the extensive and enhancer enhancers marks, we see how they overlap both the lead SNP (red dashed vertical line), and the fine-mapped Consensus SNP (vertical yellow line). This overlap is also notable on the row of Enhancers-TSS interaction marks.

**Figure 3b.**
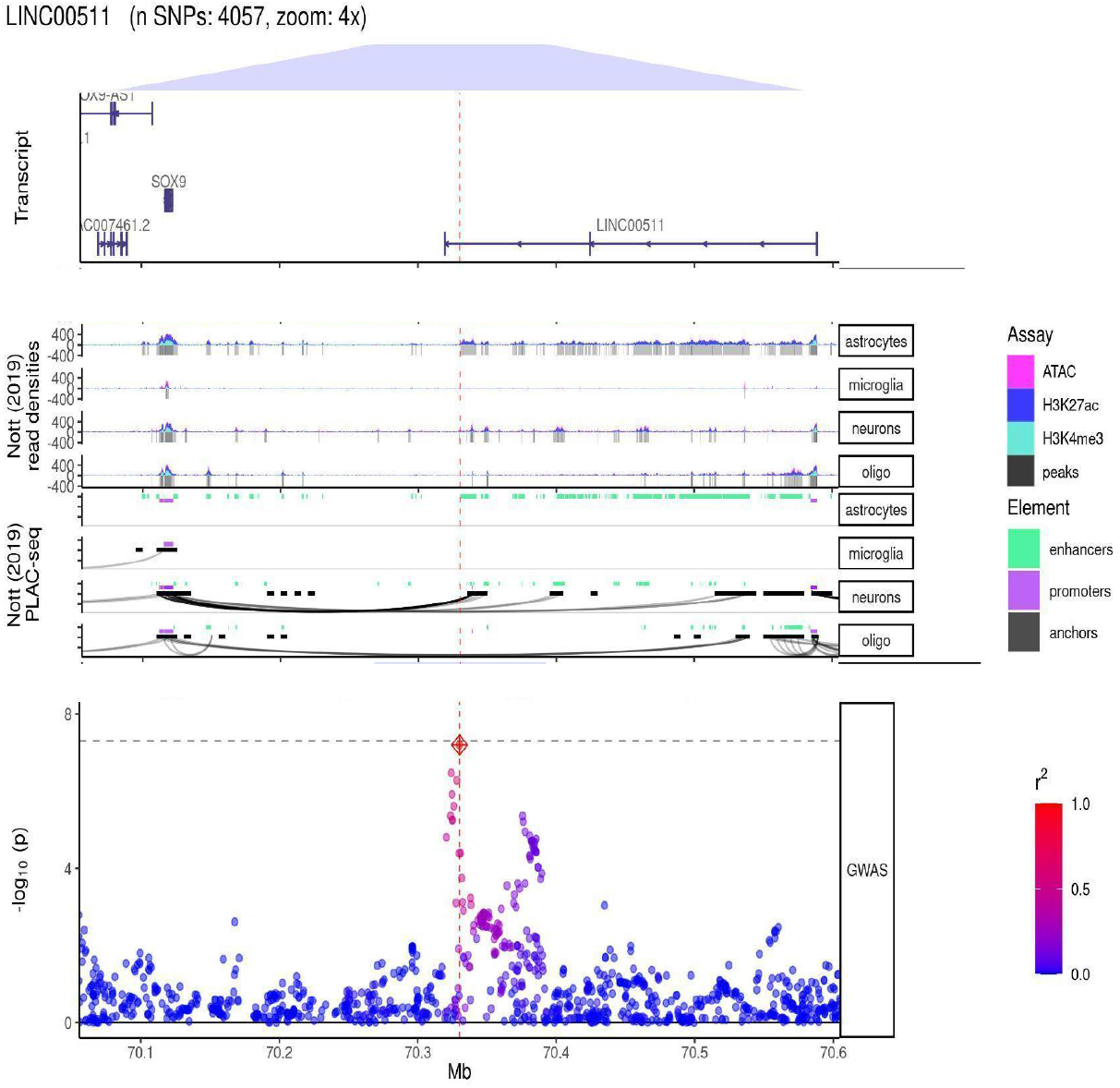
Transcripts plot (top), cell type specific regulatory element marks from Nott and colleagues (middle), locus plot (bottom). In the middle plot, the first 4 rows are the density marks (y-axis) from ATAC-seq assay (in pink), and CHIP-seq assays (H3K27ac in blus, anf H3K4me3 in cyan), in astrocytes, microglia, neurons, and oligodendrocytes. The next four rows are the distal anchored chromatin loops (black curves). We see how, only in neurons, there is a chromatin loop forming from the lncRNA towards the *SOX9* promoter.

We then performed a co-localization analysis to investigate regulation in expression of any gene controlled by the GWAS variants associated with axial motor severity and progression. We used coloc software for all genes within ±1Mb from the GWASs lead SNPs using the eQTLGen and MetaBrain Cortex tissue meta-analysis eQTL data, and PsychENCODE cortex tissue eQTL data as part of a sensitivity analysis to take into account the number of overlapping SNPs in the genes tested for colocalization (eMethods in Supplement). We did not find eQTL data for *SOX9*. We did not find evidence of colocalization at the *MAD1L1* or *GJA5* loci. In the *MAD1L1* locus, the posterior probability H3 (PPH3) (association with both phenotypic and expression traits, but distinct causal variants), was the highest (PPH3 in *MAD1L1*: eQTLGen = 0.97, MetaBrain = 0.98, PsychENCODE = 0.75). At the *GJA5* locus, we found PPH3 to be the highest for the *ACP6* gene using default SNP priors (eQTLGen = 0.98, MetaBrain = 0.88). The PPH3 remained the highest for these two genes (H3 > 0.8), after we adjusted the priors according to the number of overlapping SNPs. Considering we did not reach genome wide-significance from the GWAS in the *MAD1L1* locus, we hypothesised that the analysis may be underpowered to find co-localization support from the coloc Bayesian approach or that the regulation in expression happened in a brain tissue not tested in this study. We generated *MAD1L1* regional plots from the GWAS and cis-eQTL data from eQTLGen (Figure 3c). The observed overlap in the loci, suggests that the association with an increase in axial motor progression found in our GWAS meta-analysis, could be mediated by dysregulation in *MAD1L1* expression. However, the overlap was not as clear based on PsychEncode cis-eQTL data (eFigure 7 in Supplement).

**Figure 3c.**
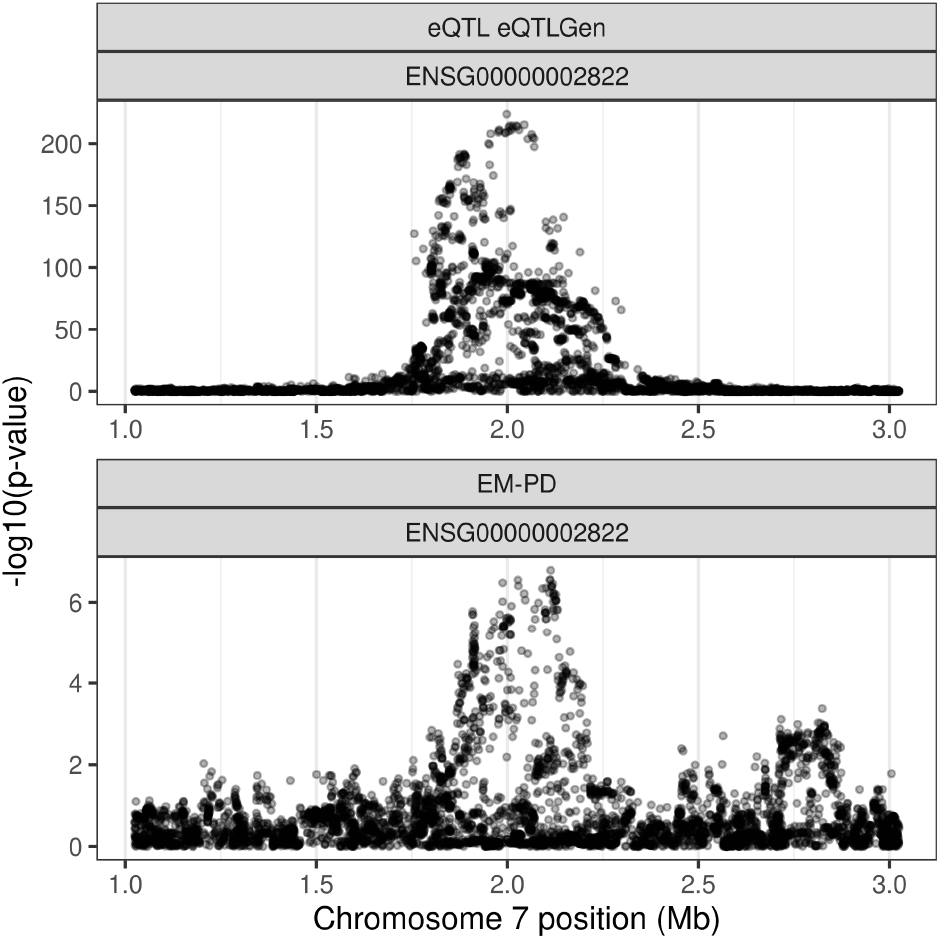
*MAD1L1* regional plots showing the overlapping variants between the axial and the eQTL trait.

Lastly, we also explored eQTLs that overlapped with the GWAS significant SNPs in FUMA. We found that many of of these signals were significant cis-eQTLs for *ACP6*, in PsychEncode, and eQTLGen (eTable 8 in Supplement). In particular, we found the lead variant to be a significant eQTL in PsychEncode (*P* = 1.2e-7), and eQTLGen (*P* = 1.7e-14), and also rs12037169, the most significant variant found in all cohorts, to be a significant cis-eQTL in eQTLGen (*P* = 8.6491e-17)

To validate the associations found with models A and B, we performed a sensitivity analysis correcting the motor scores by levodopa equivalent daily dose as described in eMethods in Supplement. We found the effect and significance to be consistent as compared to the unadjusted analysis (eTable 6 in Supplement). Hoehn and Yahr (HY), a measure of a patient’s disability, is similar to the axial score from MDS-UPDRS part III. To further validate the GWS association linked to PD axial progression, we ran model B using Hoehn and Yahr (HY), a measure of patient’s disability, as a longitudinal quantitative outcome. We found the LD block within *GJA5* nominally associated with the axial motor progression, with a consistent directionality (Beta = -0.08, SE = 0.0078, *P* = 5.7e-7). The lead variant, in the GWAS meta-analysis using HY as the outcome, rs36005900, is in LD with the lead variant reported in the MDS-UPDRS III axial GWAS (D’=0.8, R^2^=0.6) (eFigure 8 in Supplement).

## Discussion

To study the motor progression in PD, we used the MDS-UPDRS III (PD motor examination) scale, which is a sensitive measure of motor progression over time. A study of untreated *de novo* PD patients in the PPMI study, followed up for 5 years to assess the progression of MDS-UPDRS, showed a linear increase of 2.4 points per year in MDS-UPDRS part III total score ^24^. In this study, we observed a similar yearly rate of change for the total MDS-UPDRS score (2.3 points/year on average) (Table 1).

We have modelled the early stages of motor Parkinson’s using LMMs under the hypothesis that common genetic variability contributes to the motor severity and rate of change for both overall, and separate aspects of motor progression. This concept is consistent with PD subtypes studies showing a differential motor presentation and progression^25–30^, as well as limb and axial components motor PD having a different cellular and pathophysiological basis. Previous work has suggested that limb parkinsonism is broadly associated with dopaminergic dysfunction and axial symptoms may relate to cholinergic dysfunction, although other cell types, neurotransmitters and pathways may be important.

We corrected each model by AAD, and sex and PCs as confounding variables. We performed a meta-analysis to further account for between cohorts heterogeneity, as cohorts we included had different inclusion and exclusion criteria, and were either genotyped with different microarrays or whole genome sequenced. Our results are not confounded by levodopa response, as defined in our sensitivity analysis (Figure 1) (Supplementary Table 1).

In this dataset we have identified common genetic variability which determines axial, but not limb motor progression. Further functional annotation analysis allowed us to nominate *MAD1L1* and *SOX9* as candidate genes associated with motor axial severity, and *ACP6* as the gene associated with differences in the axial rate of change. Our findings were replicated across cohorts. When we explored the meta-analysis results across each study, we found that the effect was consistent across cohorts, except for PD-STAT in model A for association in the *MAD1L1* locus, and PDBP in model B.

The lack of association between common genomic variation and the MDS-UPDRS limb subscale could be due to a combination of limited power and the levodopa effect in early disease. Evers and colleagues reported that measures of mobility, tremor, gait and posture, were consistent and reliable measures of PD progression^31^. Because these measures are well represented in the axial score (except for tremor), this may be better powered to assess progression. Moreover, the limb signs may be more sensitive to levodopa use than the axial signs, making it possible that true genetic associations with limb motor progression were masked.

Overall, we have here nominated three strong candidates *MAD1L1*, S*OX9* (explaining differences in the motor axial severity), and *ACP6* (explaining differences in the axial rate of progression), supporting the concept that these elements of PD have a separate cellular and physiological basis. We have demonstrated how these genes are likely to be dysregulated in PD patients carrying the causal genetic variants by performing functional annotation. These genes can be targeted in experimental studies to further validate our findings and be considered in drug development to improve the progressive motor impairment of PD. In addition, our large scale pooled data from multiple cohorts also suggests that axial progression is a sensitive marker of change in early PD and may be a useful outcome measure in disease modifying treatment trials.

## Supporting information

EMPD

## Data Availability

All data produced are available online

https://doi.org/10.5281/zenodo.7257484

## Data availability

We have made our summary statistics available (https://doi.org/10.5281/zenodo.7257484). TPD data is available upon access request from https://www.trackingparkinsons.org.uk/about-1/data/. The PDBP and PPMI data was accessed from Accelerating Medicines Partnership: Parkinson’s Disease (AMP-PD) and data is available upon registration at https://www.amp-pd.org/. OPDC data is available upon request from the Dementias Platform UK (https://portal.dementiasplatform.uk/Apply). DIGPD data is available upon request to the principal investigator (JC Corvol, Assistance Publique Hôpitaux de Paris; https://clinicaltrials.gov/ct2/show/NCT01564992). PD-STAT is available upon request to the principal investigator (C Carroll, Plymouth University, https://penctu.psmd.plymouth.ac.uk/pdstat/#:~:text=PD%20STAT%20%2D%20Simvastatin%20as%20a,brain%20from%20injury%20or%20loss.). HapMap phase 3 data (HapMap3) is available for download at ftp://ftp.ncbi.nlm.nih.gov/hapmap/. Cis-QTL data were obtained from eQTLGen (https://www.eqtlgen.org/cis-eqtls.html) and PsychENCODE (http://resource.psychencode.org). MetaBrain cis-eQTL data can be accessed upon access request form (https://www.metabrain.nl/cis-eqtls.html). eQTL data from eQTL catalogue can be ftp-accessed (https://www.ebi.ac.uk/eqtl/Data_access/). FANTOM5 CAGE-seq and Nott brain cell type-specific enhancer-promoter interactome data were accessed through echolocatoR (https://github.com/RajLabMSSM/echolocatoR)

## Code availability

Code used in the analysis is available from https://github.com/AMCalejandro/EMPD (https://doi.org/10.5281/zenodo.7258985). Analysis was performed using open-source tools as described in the Methods section.

## Acknowledgements

This research was funded in whole or in part by Aligning Science Across Parkinson’s [Grant number: ASAP-000478] through the Michael J. Fox Foundation for Parkinson’s Research (MJFF). For the purpose of open access, the author has applied a CC BY public copyright licence to all Author Accepted Manuscripts arising from this submission. This research was supported by the National Institute for Health Research University College London Hospitals Biomedical Research Centre. The UCL Movement Disorders Centre is supported by the Edmond J. Safra Philanthropic Foundation.

Data used in the preparation of this article were obtained from the AMP-PD Knowledge Platform (https://www.amp-pd.org). AMP-PD is a public-private partnership managed by the FNIH and funded by Celgene, GSK, Michael J. Fox Foundation for Parkinson’s Research, the National Institute of Neurological Disorders and Stroke, Pfizer, and Verily. Clinical data and biosamples used in preparation of this article were obtained from the Parkinson’s Progression Markers Initiative (PPMI), and the Parkinson’s Disease Biomarkers Program (PDBP).

PPMI – a public-private partnership – is funded by the Michael J. Fox Foundation for Parkinson’s Research and funding partners, including [list the full names of all of the PPMI funding partners found at https://www.ppmi-info.org/about-ppmi/who-we-are/study-sponsors]. The PPMI Investigators have not participated in reviewing the data analysis or content of the manuscript. For up-to-date information on the study, visit www.ppmi-info.org.

The Parkinson’s Disease Biomarker Program (PDBP) consortium is supported by the National Institute of Neurological Disorders and Stroke (NINDS) at the National Institutes of Health. A full list of PDBP investigators can be found at https://pdbp.ninds.nih.gov/policy. The PDBP Investigators have not participated in reviewing the data analysis or content of the manuscript. The DIGPD cohort (ClinicalTrials.gov, NCT01564992) is a multicenter longitudinal cohort conducted in four Universities and four General Hospitals in France between 2009 and 2019, sponsored by Assistance Publique Hôpitaux de Paris, funded by a grant from the French Ministry of Health (PHRC 2008, AOR0810) and a grant from the Agence Nationale de Sécurité et des Médicaments (ANSM-2013). We thank the DIGPD Study group which collected the data made available for this work.

Both TPD and OPDC cohorts are primarily funded and supported by Parkinson’s UK (https://www.parkinsons.org.uk/) and supported by the National Institute for Health and Care Research (NIHR) Clinical Research Network (CRN). The TPD study is also supported by NHS Greater Glasgow and Clyde. The OPDC cohort is also supported by the NIHR Oxford Biomedical Research Centre, based at the Oxford University Hospitals NHS Trust, and the University of Oxford. TPD has multi-centre research ethics approval from the West of Scotland Research Ethics Committee: IRAS 70980, MREC 11/AL/0163 (ClinicalTrials.gov, NCT02881099). OPDC has multi-centre research ethics approval from the South Central Oxford A Research Ethics Committee 16/SC/0108. Each subject provided written informed consent for participation.

PD-STAT is funded and supported by grants from the Cure Parkinson’s Trust (https://cureparkinsons.org.uk/) and JP Moulton Charitable Foundation (https://www.perscitusllp.com/moulton-charity-trust/), co-ordinated by the Peninsula Clinical Trials Unit, University of Plymouth and sponsored by University Hospitals Plymouth NHS Trust. The Genotype-Tissue Expression (GTEx) Project was supported by the Common Fund of the Office of the Director of the National Institutes of Health, and by NCI, NHGRI, NHLBI, NIDA, NIMH, and NINDS. The data used for the analyses described in this manuscript were obtained from https://console.cloud.google.com/storage/browser/gtex-resources on 01/26/2022 The PsychENCODE data was generated as part of the PsychENCODE Consortium. Visit 10.7303/syn26365932 for a complete list of grants and PIs. The data was obtained from http://resource.psychencode.org/.

## Author Contributions

H.R.M. and A.M.C designed the study. H.R.M. supervised the study. D.G.G., M.T.M.H., Y.B.-S., M.A.L,, J.H. and H.R.M. conceived and led the TPD and OPDC clinical cohorts, as well as performed data management and curation. J.C.C. conceived and led the DIGPD clinical cohort, as well as performed data management and curation. A.M.C performed all the quality control and analyses in the cohorts included in the present manuscript. R.R supervised the trajectory of the research giving valuable suggestions in ways to move forward as the research progressed. C.C. led the PD STAT study, as well as performed data management and curation. M.M.X.T. helped with the design of the quality control strategy. L.W provided access to an harmonised version of the AMP-PD genetic data. R.H.R helped with the colocalization analysis and the development of regional plots. M.A.L provided the equations to adjust the motor outcomes based on levodopa usage. M.S reviewed many steps performed in the research process. M.S provided the code to perform power calculation. A.M.C wrote the initial manuscript. All authors critically reviewed the manuscript

