## Supplementary material for "Association of genetic variation at the GJA5/ACP6 locus with motor progression in Parkinson’s": EMPD

**Supplementary Online Content**

**eMethods.** Imputation of missing outcomes, Quality control and imputation, Sensitivity analysis, Statistical models evaluation, Models A and B explained, Power calculation approach, SCEBE algorithm, Software and open data for functional annotation, colocalization sensitivity analysis

**eResults.** Function of nominated genes and link to Parkinson's

**eTable 1.** Study sample sizes and genotyping array.

**eTable 2**. Breakdown of MDS-UPDRS part III scale.

**eTable 3**. Exploration in the rate of missingness of MDS-UPDRS part III across cohorts

**eTable 4**. Genomic inflation factor (λ) in each cohort across GWAS meta-analyses.

**eTable 5.** SNPs associated with axial motor score under model A and B

**eTable 6.** Breakdown of metrics for rs36082764, rs4721411, and , and rs12037169 across cohorts

**eTable 7.** Fine-mapping results of *MAD1L1* and *LINC00511* loci

**eTable 8.** Model B significant SNPs that are nominally significant cis-eQTLs

**eFigure 1**. Quality Control flowchart

**eFigure 2**. Equations predicting the impact of levodopa dosage in MDS-UPDRS part III.

**eFigure 3**. LMMs assumptions checking

**eFigure 4**. SCEBE validation in OPDC and DIGPD cohorts.

**eFigure 5**. Power to detect genetic associations from using LMMs.

**eFigure 6.** Fine Mapping and regional plots of *MAD1L1* and *LINC00511* loci.

**eFigure 7**. *MAD1L1* Regional plots from axial motor GWAS and PsychEncode cis-eQTL

**eFigure 8.**. Manhattan plot for model B GWAS meta-analysis using HY as the outcome.

**eReferences.**

#### eMethods. Decision to include data up to 3 years, Imputation of missing outcomes, Quality control and imputation, Sensitivity analysis, LMMs comparison, Models A and B explained, Power calculation approach, SCEBE algorithm, Software and open data for functional annotation, colocalization sensitivity analysis

##### 1. Decision to include data up to 3 years.

In order to study the early stages of PD, we set a threshold of three years from baseline. One reason we defined the early stages of PD as the first 3 years from the study entry, was to account for the patient's withdrawal. We found that up to three years, the missingness rate was lower than 50% in all cohorts, as well as this time visit was shared across all studies. Moreover, motor progression measured by HY and UPDRS decreases with advancing disease duration and longer follow-up, which could decrease the variability in progression as later time visits are included, hence limiting the ability to detect changes in motor scores related to genetic variability[1].

##### 2. Imputation of missing outcomes.

For participants with incomplete MDS-UPDRS part III data we scaled up the limb, axial, and total scores. For each patient's time specific MDS-UPDRS part III measures, when no more than 20% of the total scores from each motor subscore (total, limb, axial) were missing at random, we scaled up the score summing the total score across motor subscores, divided by the number of non missing subscores, multiplied by the total number of scores on each motor subscore[2] (Imputation function available on GitHub). If more than 20% of the total scores per motor subscale were missing, we set the motor subscale as missing, and excluded that data point. On the other hand, if there were items in the MDS-UPDRS part III scale consistently missing (missing not at random), we scaled up the total motor score only when there were up to 3 measures missing not at random[2].

##### 3. Quality control and imputation.

We applied standard sample QC steps across cohorts using plink PLINK v1.9 (RRID:SCR_001757; https://www.cog-genomics.org/plink/1.9/)[3].

**Sample/Patient QC**: At the patient level, we removed samples with low genotyping rates (<98%), sex mismatch between reported sex and the genotype derived sex, heterozygosity outliers (we considered samples as heterozygosity outliers if they deviated more than +-3 SD away from the mean cohort heterozygosity rate). To remove one of paired related individuals, using GCTA software (version 1.93.0 beta for Linux; https://yanglab.westlake.edu.cn/software/gcta/#Overview)[4], we created a genomic relationship matrix from pruned data, and individuals with a SNP similarity (PIHAT) higher than 0.0875, equivalent to 1st degree relative, were excluded. To deal with population stratification, we performed a principal component analysis (PCA) over pruned genotype data of each independent cohort merged with Utah residents with Northern and Western European ancestry (CEU), Han Chinese in Beijing, China (CHB), Japanese in Tokyo, Japan (JPT), and Yoruba in Ibadan, Nigeria (YRI) populations from the HapMap reference penel to identify non-European ancestry sample [5]. At first, we visualised each cohort with CEU, CHB, JPT, and YRI populations, so as to make a decision on the threshold of standard deviations (SD) away from any of the mean 10 first PCs from the CEU population to consider non-European ancestry samples. Finally, a second filter was applied to further remove heterozygosity outliers, as well as samples with low genotyping rate (<95%) based on recalculated relatedness and missingness frequencies on the remaining samples.

**Variant QC**: At a genotype level, we removed variants that had a missing rate higher than 0.05, variants with a minor allele frequency (MAF) of less than 0.01, and variants whose missing calls were not randomly distributed by testing whether missingness status could be predicted from genotype calls at the two adjacent variants. Moreover, we excluded variants that were deviated from the Hardy-Weinberg equilibrium (HWE) as extreme HWE deviations are indicative of sample contamination. (P Value<1e-10) [6].

**Imputation**. After QC, we individually imputed all the chip genotyped cohorts. Imputation is a method to increase the number of variants available to test in genome wide studies. This method matches genotype chip data haplotypes to Whole Genome Reference haplotypes, then inferring the complete haplotype from the full Reference Panel Haplotype. In order to prepare data for imputation, we ran the Will Rayner tool for further quality checks according to the Haplotype Reference Consortium (HRC) Panel (version r1.1 2016; http://www.haplotype-reference-consortium.org/). Prior imputation, we updated strand, position, and reference / alternate allele assignment, as well removing A/T and G/C SNPs if MAF> 0.4, SNPs with > 0.2 allele frequency difference, and SNPs not present in the HRC Panel [7]. Then, we imputed in the Michigan Imputation Server (MIS), using Minimac4 (version 1.0.0, released 2.14.2018; https://genome.sph.umich.edu/wiki/Minimac4 version 1.0.0; RRID:SCR_009292) as the genotype imputation software, HRC as the Reference Panel for imputation, and Eagle v2.4 (RRID:SCR_017262; https://alkesgroup.broadinstitute.org/Eagle/) for phasing out.

**Post-imputation QC**: To only work with variants that were imputed with high confidence, we removed those with an Rsq < 0.8. Furthermore, we excluded variants with low genotyping rate (<95%), and MAF < 0.01, resulting in over 500000 SNPs available across cohorts (Supplementary Figure 3).

**Post-meta-analysis QC:** Once the meta-analysis was complete, we removed variants with MAF variability higher than 15%, and also those variants showing high between-study heterogeneity according to the Cochran's Q-test ($P < 0.05$) and $I^2$ index (variants with an heterogeneity higher than 80%).

##### 4. Sensitivity analysis

To take into account the effect of dopaminergic treatment and doses on the motor scores derived from MDS-UPDRS part III, we performed a sensitivity analysis with the adjusted total, limb, and axial motor scores, using a correction factor according to the effect of levodopa dose on the MDS-UPDRS scale.

It is well known that levodopa treatment slows disease progression in the majority of PD patients [8]. To figure out whether any genetic association with the motor states was masked due to levodopa dosage, we used an equation that best predicted the effect of levodopa dose on MDS-UPDRS part III total over time to correct the motor scores by levodopa usage, provided by Dr Michael Lawton at University of Bristol (eFigure 2). We used data from Tracking Parkinson's Levodopa challenge with motor subscores recorded at baseline before and after treatment in order to weight the effect of levodopa usage on the limb and axial motor states. We found that over the average difference in the MDS-UPDRS part III total score pre and post dose at baseline (9.9 points difference in average), 7.7 point change was explained by the limb composite score and 2.2 change due to the axial score. We used such weights on the equation that best predicted what we would expect to happen long term with levodopa usage, and we derived the adjusted outcomes each time visit across cohorts.

##### 5. LMMs comparison

We carried out a comparison of different LMMs by using BIC, and marginal and conditional R Squared metrics based on patient's data from the TPD cohort. To avoid overfitting, BIC introduces a penalty parameter depending on the number of covariates added to the model, allowing us to select the model that explains the highest percentage of variance with the lowest combination of predictors. The lower the BIC, the better. The marginal R Squared shows the variance explained by the conditional likelihood and the conditional R Squared shows the variance explained from both random and fixed parts of the model.

We adjusted all models by gender, age at diagnosis (AAD), and PCs derived from pruned genetic data to account for any remaining population structure. Then, we compared LMMs fitted with different combinations of predictors. We did not find a decrease in BIC after inclusion of any covariate as a fixed effect, except for a measure of the motor score at baseline. However, this model did not meet the LMMs linearity assumptions, so we discarded including it as a predictor in the model. When we added time random slopes to the model adjusted by AAD and the confounding variables, we noticed a decrease in BIC. In addition, the conditional R squared increased significantly.

##### 6. Models A and B explained.

We acknowledge that PD is a heterogeneous disorder [9–11]. Nevertheless, another undesired and realistic contributor to this high heterogeneity found when studying different PD cohorts could be linked to events such as patien misdiagnosis, as well as the underlying noise that MDS-UPDRS part III scale has [12]. Therefore, we decided to fit LMMs based on two principles. We used model A to capture genetic determinants explaining differences in the average patients motor scores, assuming there is no unexplained variability at the slope level, and model B to capture genetic variability associated with a faster or slower rate of progression, which is the model with the most favourable (lowest) BIC while meeting the LMMs assumptions.

**(A)** OUTCOME ~ 1 + (1|ID) + SNP + GENDER + AGE_DIAGNOSIS.STD + 5PCs

**(B)** OUTCOME ~ 1 + (1 + TIME |ID) + SNP + TIME*SNP + GENDER + AGE_DIAGNOSIS.STD + 5PCs

Model (A) is a LMM with random variability at the intercept level only. We allow the individual's intercepts to deviate from the global intercept. It is adjusted by the confounding variables only. We selected model (A) under the assumption there is no unexplained variability in Parkinson's disease progression

Model (B) is a LMM with random variability at both the intercept and the slope level. We allowed for individual's intercepts to deviate from the global intercept as well as time individual's slopes to deviate from the global average time slope, while allowing correlation between the intercept deviations and time effect deviations within individual levels. We selected model (B) under the assumption that there are differences in patients' progression that could be explained from genetics.

##### 7. Power calculation approach

The power of GWASs depends on the sample size, the frequency of those variants associated with traits, their effect size, the heterogeneity of the trait studied, and the LD structure. As the sample size of the population studied, the effect of a trait-associated variant, and the allele frequency increases, the power to genetic associations increases. Interestingly, it is worth noting that there is not much difference in power when doing GWAS based on imputed data from SNP arrays or WGS except for ultra-rare variants (1e-5) [13]. To estimate the power of LMMs in GWAS, we performed a power calculation across combinations of sample sizes, allele frequencies, and effect sizes in R. We carried out 10000 simulations and tested the association of 1000 dummy SNPs with different effect sizes (total MDS-UPDRS III rate of decline), different AFs for those SNPs, and for three different sample sizes.We reported the power as the number of times a SNP was found to be significantly associated with the outcome accounting for multiple testing ($P$ = 0.05 / N SNPs), divided by the number of simulations.

##### 8. SCEBE algorithm

When using some type of more complex statistical models under a GWAS setting, the job can become computationally expensive. An example is when we use LMMs allowing for random variability at both the intercept and the slope level, with covariates as well as interaction terms. SCEBE is an algorithm adapted to explore genome-wide associations with longitudinal outcomes through mixed-effect modelling. With SCEBE, we fitted a base mixed-effects model with REML and we used the predictors of random effects from the base model as phenotypes for GWAS through a simple linear regression model. Because the predictors of random effects are affected from the shrinkage to population mean as they are the weighted sum of the population and sample mean, using them as phenotypes would lead to biassed estimations of the SNP effect estimated ad P-values. Yuan and colleagues quantified in SCEBE the bias and added it as a correction matrix, allowing us to generate unbiased SNPs test statistics [14].

##### 9. Software and open data for functional annotation

We used CGTA-COJO software (version 1.93.0 beta for Linux; https://yanglab.westlake.edu.cn/software/gcta/#mtCOJO) to find the number of independent mutations associated with PD motor phenotypes[15]. To perform fine-mapping we used coloc (version 5.1.0; https://cloud.r-project.org/web/packages/coloc/index.html), FINEMAP (v.1.3.1; http://www.christianbenner.com/), and SuSiE (v. 0.12.27; https://github.com/stephenslab/susieR), statistical fine-mapping methods, and PolyFun-SuSiE (v. 1.0; https://github.com/omerwe/polyfun) [16–19]. We used UK Biobank (https://www.ukbiobank.ac.uk/; RRID:SCR_012815)[20] as a reference panel to retrieve the loci LD structure necessary to compute SNPs posterior probabilities from GWAS summary statistics. We set to 95% the fine-mapped posterior probability for a SNP to be considered part of a Credible Set ($CS_{95\%}$). Therefore, each CS represents the minimum set of SNPs that contains the causal SNP with probability 95%. We used echolocatoR R package (V. 1.0; https://github.com/RajLabMSSM/echolocatoR) to report the Union Credible Set SNPs (UCS), which is the union of all tool-specific $CS_{95\%}$, as well as the Consensus SNPs, which are those nominated from at least two fine mapping tools. Moreover, we performed colocalization analysis of the EM-PD GWASs against cis-eQTL data made available through eQTL Catalogue, a repository of uniformly processed gene expression and splicing QTLs from all available public studies on human (https://www.ebi.ac.uk/eqtl/, accessed on the 10/07/2022)[21], as well as PsychEncode, comprising gene expression from bulk RNA sequencing from the cerebral neocortex of 1,387 individuals (http://resource.psychencode.org/; accessed on the 10/07/2022), eQTLGen, comprising bulk blood-derived gene expression from 31,684 individuals (https://www.eqtlgen.org/cis-eqtls.html, accessed on the 10/07/2022), and MetaBrain cis-eQTL, large scale eQTL meta-analysis of previously published human brain tissues eQTL datasets (https://www.metabrain.nl/; accessed on the 1/08/2022) resources[22–24]. To perform co-localization in loci between GWAS and cis-eQTL datasets, we used the colochepR R package (v. 0.99.0)[25] as a wrapper. We used echolocatoR to access the Nott and colleagues and FANTOM5 regulatory elements datasets named in the main text.

#### 10. Colocalization sensitivity analysis

We used coloc software to test colocalization for all genes within ±1Mb from the GWASs lead SNPs using the eQTLGen and MetaBrain Cortex tissue meta-analysis eQTL data. We used these two datasets as they are the largest blood and brain eQTL studies respectively, providing us with the greatest power to perform statistical co-localization tests. However, it is worth noting that the prior for H3 hypothesis (association with both phenotypic and expression traits, but distinct causal variants) is $\approx n(n-1)p1\ p2$, which scales with the square of n, resulting in H3 becoming more likely than H4 as the number of overlapping SNPs in the region tested increases [26]. This affects the colocalization tests against MetaBrain and eQTLGen meta-analyses. Therefore, we performed two sensitivity analyses, adjusting the priors according to the number of overlapping SNPs[27], and also performing co-localization against PsychENCODE , the largest cortex tissue eQTL dataset from a single study, which resulted in a considerable decrease in overlapping SNPs compared to the overlap against eQTL meta-analyses.

### eResults. Function of nominated genes and link to Parkinson's.

#### 1. Function of nominated genes and link to Parkinson's

*MAD1L1* encodes the mitosis arrest deficient-like 1 protein, a component of the spindle-assembly checkpoint which prevents the onset of anaphase until chromosomes are aligned at the metaphase plate [28]. Recent GWAS have identified *MAD1L1* as a gene increasing the susceptibility for bipolar disorder and schizophrenia [29,30]. This variant is in high LD with the fine-mapping *MAD1L1* nominated variant (D' = 0.75) [31]. *MAD1L1* expression is measurable in several brain tissues [32]. A recent study investigated healthy adults carrying the *MAD1L1* rs11764590 risk allele[33]. Carriers showed alteration in the responsiveness and regulation of the mesolimbic reward system. Adults carrying the risk alleles showed significant hypoactivations of the ventral tegmental area (VTA), the bilateral striatum, and bilateral frontal and parietal cortices. Regarding PD in particular, a study including PD patients has shown that patients with more severe disease (measured in "OFF" and "ON " state), showed a fall in activation in the anterior cingulate cortex associated with reward expectancy[34]. A plausible explanation for this could be that *MAD1L1* PD mutation carriers, showing an impaired reward system, respond worse to dopaminergic therapy, hence developing with more severe axial signs.

It is known that enhancers are found in intronic and intergenic regions, as well as that introns act as gene regulators [35,36]. We have found evidence of the *MAD1L1* intron acting as an active enhancer and regulating and predicted to interact with a transcription start site (TSS). This together with the overlap found between eQTL and GWAS *MAD1L1* regional plots, suggest that this intron may play an active role in regulation in expression.

*SOX9* is a SOX transcription factor (TFs) family member. The male sex determination gene (Sry) gave birth to this SOX family. SOX TFs regulate diverse cellular processes during development, as well as differentiation into tissues and organs. In addition, they play a major role in central nervous system development and adult neurogenesis[37]. Studies of *SOX9* gain and loss of function have demonstrated that *SOX9* is required for the formation of multipotent Neural stem cells (NSCs) and their maintenance in the central nervous system during embryonic and adult phase[38]. Moreover, *SOX9* regulates the transition from neurogenesis to gliogenesis during development, and it has been shown that when *SOX9* is not expressed, there was a reduction in astrocytes and oligodendrocytes, and a transient increase in motor neurons[39,40]. This is consistent with our findings suggesting when the distal regulation towards *SOX9* expression is altered in neurons, PD patients show a lower motor axial impairment, suggesting a connection between the CNS development and the adult neurogenesis.

*GJA5* is a member of the connexin gene family. The connexin coded by *GJA5* is CX40, which is a component of gap junctions in astrocytes and neurons which enable the signalling between the cytoplasm of adjacent cells. Gap junctions form the neuron-glial network and are key for the maintenance of homeostasis[41]. In PD in particular, an enhanced expression of the related gene *GJA1*, a gene coding the connexin CX43 has been found in the striatum of rat models and cultured astrocytes with rotenone[42]. However, the link between an increased gap junction intercellular communication (GJIC) and dopaminergic cell death is still unknown[43]. Another study found in mouse cortical astrocytes that α-synuclein enhanced the opening of CX43 hemichannels, suggesting that the regulation of such channels may play a role in the pathogenesis and procession of synucleinopathies. Therefore, we hypothesised that this particular mutation could be protective for PD through a reduction in the expression levels, although no gene expression data was available to prove this.

We also found *ACP6* as a strong gene candidate nearby *ACP6* (distance from *GJA5* lead SNP < 1Mb). *ACP6* is an enzyme that regulates lipid metabolism in mitochondria[44]. *ACP6* irregular concentrations are found in Gaucher Disease (GD), although there is no clear link between *ACP6* levels in and GD progression. *ACP6* has a high astrocytes specificity[45]. Mitochondrial dysfunction has been widely associated with PD aetiology [46]

#### eTable 1. Study sample sizes and genotyping array.

| Study Name | Samples source | Abbreviations | N | Genotyping array |
|---|---|---|---|---|
| Tracking Parkinson's | UK | TPD | 2000 | Illumina HumanCoreExome array |
| Oxford Parkinson's Disease Centre Discovery Cohort | UK | OPDC | 1082 | Illumina HumanCoreExome-12 v1.1 or Illumina Infinium HumanCoreExome-24 v1.1 |
| Drug Interaction With Genes in Parkinson's Disease | FRANCE | DIGPD | 427 | umina Infinium Multi-Ethnic Global (MEGA) |
| Parkinson's Progression Markers Initiative | US | PPMI | 415 | WGS |
| Advancing Parkinson's Disease Biomarkers Discovery | US | PDBP | 873 | WGS |
| Simvastatin as a neuroprotective treatment for Parkinson's disease | UK | PD-STAT | 174 | Neurochip |

WGS = Whole Genome Sequenced

#### eTable 2. Breakdown of MDS-UPDRS part III scale

| Motor Score | Scores from MDS-UPDRS III |
|---|---|
| MDS-UPDRS part III - Total | Speech (3.1) , Facial expression (3.2), Rigidity (3.3), Finger tapping (3.4), Hand movement (3.5), Pronation-supination movements of hands (3.6) , toe tapping (3.7), leg agility (3.8), Arising from chair (3.9), Gait (3.10), freezing of gait (3.11), postural stability (3.12), Posture (3.13) global spontaneity of movement body (Body bradykinesia) (3.14), postural tremor of the hands ( 3.15), kinetic tremor of the hands (3.16), rest tremor amplitude (3.17), constancy of rest tremor (3.18) |
| MDS-UPDRS part III - Limb | Rigidity (3.3), postural tremor of the hands (3.15), kinetic tremor of the hands (3.16), rest tremor amplitude (3.17) Finger tapping (3.4), Hand movement (3.5), Pronation-supination movements of hands (3.6) , toe tapping (3.7), leg agility (3.8), constancy of tremor (3.18) |
| MDS-UPDRS part III - Axial | Speech (3.1) , Facial expression (3.2), Arising from chair (3.9), Gait (3.10), freezing of gait (3.11), postural stability (3.12), Posture (3.13), global spontaneity of movement body (Body bradykinesia) (3.14) |

#### eTable 3. Exploration in the rate of missingness of MDS-UPDRS part III across cohorts

| Cohort | Missingness before Imputation | | | | | Missingness after Imputation | | | | |
|---|---|---|---|---|---|---|---|---|---|---|
| | Baseline | 12 months | 18 months | 24 months | 36 months | Baseline | 12 months | 18 months | 24 months | 36 months |
| TPD | 9.60 | X | 0.24 | X | 0.39 | 0.01 | X | 0.14 | X | 0.29 |
| OPDC | 0.02 | X | 0.23 | X | 0.37 | 0.01 | X | 0.18 | X | 0.35 |
| PPMI | 0.00 | 0.28 | 0.51 | 0.38 | 0.39 | 0.00 | 0.28 | 0.51 | 0.38 | 0.39 |
| DIGPD | 0.00 | 0.08 | X | 0.15 | 0.28 | 0.00 | 0.08 | X | 0.15 | 1.28 |
| PD-STAT | 0.00 | 0.03 | X | 0.13 | X | 0.00 | 0.01 | X | 0.09 | X |
| PDBP | 0.00 | 0.09 | 0.20 | 0.23 | 0.39 | 0.00 | 0.07 | 0.18 | 0.22 | 0.36 |

#### eTable4. Genomic inflation factor (λ) in each cohort across GWAS meta-analyses.

| | model A | | | LEDD adjusted model A | | | model B | | | LEDD adjusted model B | | |
|---|---|---|---|---|---|---|---|---|---|---|---|---|
| | total | limb | axial | total | limb | axial | total | limb | axial | total | limb | axial |
| TPD | 1.01 | 1.01 | 1.01 | 1.01 | 1.02 | 1.01 | 1.00 | 1.01 | 1.00 | 1.00 | 1.00 | 1.00 |
| OPDC | 1.02 | 1.01 | 1.01 | 1.02 | 1.02 | 1.02 | 1.00 | 1.00 | 1.01 | 1.00 | 1.00 | 1.00 |
| PPMI | 1.04 | 1.04 | 1.04 | 1.05 | 1.05 | 1.05 | 0.99 | 1.00 | 1.00 | 1.00 | 1.00 | 1.00 |
| DIGPD | 1.10 | 1.10 | 1.11 | 1.05 | 1.05 | 1.05 | 1.02 | 1.05 | 1.19 | 0.99 | 0.99 | 0.97 |
| PDSTAT | 1.05 | 1.04 | 1.05 | NA | NA | NA | 0.99 | 0.99 | 0.98 | NA | NA | NA |
| PDBP | 1.00 | 1.00 | 1.00 | NA | NA | NA | 0.99 | 1.00 | 0.99 | 0.99 | 1.00 | 1.00 |

NA = Cohort was not included in the analysis due to missing data needed.

#### eTable 5. SNPs associated with axial motor score under model A and B

| rsID | chr | pos | A1 | A2 | MAF | Beta | se | P-value | nearest Gene | distance | type of variant | Model |
|---|---|---|---|---|---|---|---|---|---|---|---|---|
| rs36082764 | 17 | 70330179 | T | C | 0.42 | -0.62 | 0.11 | 6.34e-08 | *LINC00511* | 0 | ncRNA_intronic | model A |
| rs4721411 | 7 | 2153071 | T | C | 0.40 | 0.53 | 0.10 | 1.66e-07 | *MAD1L1* | 0 | intronic | model A |
| rs10939702 | 4 | 10096692 | T | G | 0.45 | 0.57 | 0.12 | 8.10e-07 | *WDR1* | 0 | intronic | model A |
| rs6593808 | 1 | 147219250 | A | G | 0.23 | -0.28 | 0.04 | 1.35e-10 | *GJA5* | 0 | intergenic | model B |
| rs12037169 | 1 | 147248057 | A | G | 0.25 | -0.25 | 0.04 | 3.93e-10 | *GJA5* | 0 | intergenic | model B |
| rs4073509 | 2 | 192611013 | C | T | 0.02 | 0.52 | 0.10 | 2.12e-07 | *AC098872.3* | 47137 | intergenic | model B |
| rs117239007 | 13 | 30550016 | C | T | 0.01 | 0.68 | 0.14 | 4.71e-07 | *LINC00544* | 25390 | intergenic | model B |

**eTable 6. Breakdown of metrics for rs36082764, rs4721411, and rs12037169 across cohorts and models**

| Cohort | rsID | Effect allele | maf | Beta | se | P-Value | Levodopa Adjusted | Model |
|---|---|---|---|---|---|---|---|---|
| TPD | rs36082764 | T | 0.44 | -0.66 | 0.17 | 2.50e-04 | No | Model A |
| OPDC | rs36082764 | NA | NA | NA | NA | NA | No | Model A |
| PPMI | rs36082764 | T | 0.46 | -0.66 | 0.19 | 6.50e-04 | No | Model A |
| DIGPD | rs36082764 | T | 0.38 | -0.26 | 0.36 | 0.44 | No | Model A |
| PDBP | rs36082764 | T | 0.43 | -0.65 | 0.28 | 1.00e-02 | No | Model A |
| PD-STAT | rs36082764 | NA | NA | NA | NA | NA | No | Model A |
| TPD | rs4721411 | T | 0.42 | 0.62 | 0.18 | 6.00e-04 | No | Model A |
| OPDC | rs4721411 | T | 0.41 | 0.65 | 0.24 | 8.00e-03 | No | Model A |
| PPMI | rs4721411 | T | 0.38 | 0.43 | 0.19 | 2.00e-02 | No | Model A |
| DIGPD | rs4721411 | T | 0.43 | 0.42 | 0.36 | 0.2 | No | Model A |
| PDBP | rs4721411 | T | 0.40 | 0.70 | 0.27 | 9.00e-03 | No | Model A |
| PD-STAT | rs4721411 | T | 0.42 | -0.40 | 0.52 | 0.45 | No | Model A |
| TPD | rs36082764 | T | 0.44 | -0.65 | 0.18 | 3.00e-04 | Yes | Model A |
| OPDC | rs36082764 | NA | NA | NA | NA | NA | Yes | Model A |
| PPMI | rs36082764 | T | 0.46 | -0.70 | 0.21 | 8.00e-04 | Yes | Model A |
| DIGPD | rs36082764 | T | 0.38 | -0.32 | 0.36 | 0.38 | Yes | Model A |
| TPD | rs4721411 | T | 0.42 | 0.62 | 0.18 | 6.00e-04 | Yes | Model A |
| OPDC | rs4721411 | T | 0.41 | 0.65 | 0.24 | 8.00e-03 | Yes | Model A |
| PPMI | rs4721411 | T | 0.38 | 0.33 | 0.21 | 9.00e-02 | Yes | Model A |
| DIGPD | rs4721411 | T | 0.42 | 0.38 | 0.36 | 0.29 | Yes | Model A |
| TPD | rs12037169 | A | 0.24 | -0.30 | 0.06 | 1.67e-06 | No | Model B |
| OPDC | rs12037169 | A | 0.25 | -0.33 | 0.09 | 1.75e-04 | No | Model B |
| PPMI | rs12037169 | A | 0.27 | -0.10 | 0.35 | 0.44 | No | Model B |
| DIGPD | rs12037169 | A | 0.24 | -0.29 | 0.11 | 1.84e-03 | No | Model B |
| PDSTAT | rs12037169 | A | 0.25 | -0.33 | 0.25 | 0.18 | No | Model B |
| PDBP | rs12037169 | A | 0.23 | 0.07 | 0.18 | 0.59 | No | Model B |
| TPD | rs12037169 | A | 0.24 | -0.30 | 0.07 | 8.41e-06 | Yes | Model B |
| OPDC | rs12037169 | A | 0.25 | -0.35 | 0.09 | 6.20e-04 | Yes | Model B |
| PPMI | rs12037169 | A | 0.27 | -0.21 | 0.39 | 0.6 | Yes | Model B |
| DIGPD | rs12037169 | A | 0.24 | -0.28 | 0.11 | 1.84e-03 | Yes | Model B |
| PDBP | rs12037169 | A | 0.23 | -0.05 | 0.13 | 0.67 | Yes | Model B |

NA = SNP missing in such cohort for the model given in Model column.

#### eTable 7. Fine-mapping results of MAD1L1 and LINC00511 loci

| SNP | CHR | N | t_stat | ABF.CS | ABF.PP | FINEMAP.CS | FINEMAP.PP | SUSIE.CS | SUSIE.PP | POLYFUN_SUSIE.CS | POLYFUN_SUSIE.PP | Support | Consensus | mean.PP |
|---|---|---|---|---|---|---|---|---|---|---|---|---|---|---|
| rs3778978 | 7 | 3500 | -4.99 | 0.00 | 0.02 | NA | NA | 3.00 | 0.96 | 3.00 | 1.00 | 2.00 | TRUE | 0.50 |
| rs7213651 | 17 | 3500 | 4.71 | 0.00 | 0.02 | 1 | 0.86 | 4.00 | 1.00 | 4.00 | 1.00 | 3.00 | TRUE | 0.72 |
| rs7218929 | 17 | 3500 | 4.54 | 0.00 | 0.01 | 3 | 0.07 | 1.00 | 1.00 | 2.00 | 1.00 | 3.00 | TRUE | 0.52 |
| rs12950478 | 17 | 3500 | -4.59 | 0.00 | 0.01 | NA | NA | 2.00 | 1.00 | 3.00 | 1.00 | 2.00 | TRUE | 0.50 |

Abbreviation = N, Sample size to do fine-mapping; t_stat = test statistic; CS = Credible Set; PP = Posterior Probability. mean.PP = the mean posterior probability from the four fine-mapping posterior probabilities

#### eTable 8. Model B significant SNPs that are nominally significant cis-eQTLs

| SNP | CHR | POS | MAF | nearestGene | gwasP | tissue | symbol | eqtlP |
|---|---|---|---|---|---|---|---|---|
| rs6593808 | 1 | 147219250 | 0.23 | *GJA5* | 1.349e-10 | PsychENCODE_eQTLs | *ACP6* | 1.18e-07 |
| rs6593808 | 1 | 147219250 | 0.23 | *GJA5* | 1.349e-10 | eQTLGen_cis_eQTLs | *ACP6* | 1.68e-14 |
| rs2353 | 1 | 147222372 | 0.23 | *GJA5* | 1.349e-10 | eQTLGen_cis_eQTLs | *ACP6* | 1.58e-14 |
| rs7551148 | 1 | 147289707 | 0.25 | *RP11-314N2.2* | 1.815e-10 | PsychENCODE_eQTLs | *ACP6* | 1.23e-07 |
| rs7551148 | 1 | 147289707 | 0.25 | *RP11-314N2.2* | 1.815e-10 | PsychENCODE_eQTLs | *LINC01138* | 1.43e-4 |
| rs7551148 | 1 | 147289707 | 0.25 | *RP11-314N2.2* | 1.815e-10 | eQTLGen_cis_eQTLs | *ACP6* | 1.08e-07 |
| rs1495955 | 1 | 147249285 | 0.25 | *RP11-433J22.3* | 2.992e-10 | PsychENCODE_eQTLs | *ACP6* | 7.97e-05 |
| rs1495955 | 1 | 147249285 | 0.25 | *RP11-433J22.3* | 2.992e-10 | eQTLGen_cis_eQTLs | *ACP6* | 6.52e-17 |
| rs12037169 | 1 | 147248057 | 0.25 | *GJA5* | 3.93e-10 | eQTLGen_cis_eQTLs | *ACP6* | 8.64e-17 |
| rs1857213 | 1 | 147219553 | 0.23 | *GJA5* | 6.383e-10 | PsychENCODE_eQTLs | *ACP6* | 1.18e-07 |
| rs1857213 | 1 | 147219553 | 0.23 | *GJA5* | 6.383e-10 | eQTLGen_cis_eQTLs | *ACP6* | 1.73e-14 |
| rs1342711 | 1 | 147219835 | 0.23 | *GJA5* | 6.383e-10 | PsychENCODE_eQTLs | *ACP6* | 1.20e-07 |
| rs1342711 | 1 | 147219835 | 0.23 | *GJA5* | 6.383e-10 | eQTLGen_cis_eQTLs | *ACP6* | 2.10e-14 |
| rs12032789 | 1 | 147220045 | 0.23 | *GJA5* | 6.383e-10 | PsychENCODE_eQTLs | *ACP6* | 7.91e-09 |
| rs12032789 | 1 | 147220045 | 0.23 | *GJA5* | 6.383e-10 | eQTLGen_cis_eQTLs | *ACP6* | 1.55e-14 |
| rs36005900 | 1 | 147229662 | 0.23 | *GJA5* | 6.964e-10 | eQTLGen_cis_eQTLs | *ACP6* | 1.92e-13 |
| rs2352870 | 1 | 147206521 | 0.26 | *GJA5* | 7.019e-10 | PsychENCODE_eQTLs | *ACP6* | 1.18e-07 |
| rs2352870 | 1 | 147206521 | 0.26 | *GJA5* | 7.019e-10 | eQTLGen_cis_eQTLs | *ACP6* | 3.01e-08 |
| rs10793706 | 1 | 147243753 | 0.25 | *GJA5* | 8.465e-10 | PsychENCODE_eQTLs | *ACP6* | 5.93e-05 |
| rs10793706 | 1 | 147243753 | 0.25 | *GJA5* | 8.465e-10 | eQTLGen_cis_eQTLs | *ACP6* | 9.65e-17 |
| rs10793707 | 1 | 147244022 | 0.25 | *GJA5* | 8.465e-10 | PsychENCODE_eQTLs | *ACP6* | 5.12e-05 |
| rs10793707 | 1 | 147244022 | 0.25 | *GJA5* | 8.465e-10 | eQTLGen_cis_eQTLs | *ACP6* | 1.50e-16 |
| rs12408247 | 1 | 147244441 | 0.25 | *GJA5* | 8.465e-10 | eQTLGen_cis_eQTLs | *ACP6* | 1.18e-16 |
| rs1692141 | 1 | 147230217 | 0.24 | *GJA5* | 9.066e-10 | eQTLGen_cis_eQTLs | *ACP6* | 1.03e-14 |
| rs1891498 | 1 | 147226791 | 0.23 | *GJA5* | 1.048e-09 | eQTLGen_cis_eQTLs | *ACP6* | 6.54e-13 |
| rs1043806 | 1 | 147229299 | 0.24 | *GJA5* | 1.048e-09 | eQTLGen_cis_eQTLs | *ACP6* | 1.34e-14 |
| rs11552588 | 1 | 147245383 | 0.25 | *GJA5* | 1.102e-09 | eQTLGen_cis_eQTLs | *ACP6* | 2.02e-16 |
| rs35594137 | 1 | 147245497 | 0.25 | *GJA5* | 1.102e-09 | PsychENCODE_eQTLs | *ACP6* | 4.94e-05 |
| rs35594137 | 1 | 147245497 | 0.25 | *GJA5* | 1.102e-09 | eQTLGen_cis_eQTLs | *ACP6* | 2.46e-16 |
| rs12408178 | 1 | 147246903 | 0.25 | *GJA5* | 1.106e-09 | eQTLGen_cis_eQTLs | *ACP6* | 4.01e-16 |
| rs6593806 | 1 | 147213538 | 0.23 | *GJA5* | 1.169e-09 | eQTLGen_cis_eQTLs | *ACP6* | 7.86e-15 |
| rs2352868 | 1 | 147210965 | 0.26 | *GJA5* | 1.197e-09 | eQTLGen_cis_eQTLs | *ACP6* | 8.51e-08 |
| rs4950344 | 1 | 147227066 | 0.23 | *GJA5* | 1.435e-09 | eQTLGen_cis_eQTLs | *ACP6* | 5.63e-13 |
| rs6669493 | 1 | 147202308 | 0.26 | *GJA5* | 3.035e-09 | PsychENCODE_eQTLs | *ACP6* | 1.30e-07 |
| rs6669493 | 1 | 147202308 | 0.26 | *GJA5* | 3.035e-09 | eQTLGen_cis_eQTLs | *ACP6* | 1.13e-07 |
| rs10900394 | 1 | 147202386 | 0.26 | *GJA5* | 3.035e-09 | PsychENCODE_eQTLs | *ACP6* | 1.31e-07 |
| rs10900394 | 1 | 147202386 | 0.26 | *GJA5* | 3.035e-09 | eQTLGen_cis_eQTLs | *ACP6* | 1.05e-07 |
| rs4412645 | 1 | 147206859 | 0.27 | *GJA5* | 4.553e-09 | PsychENCODE_eQTLs | *ACP6* | 4.16e-07 |
| rs1891504 | 1 | 147209539 | 0.26 | *GJA5* | 5.353e-09 | PsychENCODE_eQTLs | *ACP6* | 2.47e-07 |

| SNP | CHR | POS | MAF | nearestGene | gwasP | tissue | symbol | eqtlP |
|---|---|---|---|---|---|---|---|---|
| rs1891504 | 1 | 147209539 | 0.26 | *GJA5* | 5.353e-09 | eQTLGen_cis_eQTLs | *ACP6* | 1.34e-07 |
| rs4469765 | 1 | 147207012 | 0.27 | *GJA5* | 5.371e-09 | PsychENCODE_eQTLs | *ACP6* | 4.16e-07 |
| rs1891505 | 1 | 147208624 | 0.26 | *GJA5* | 5.417e-09 | PsychENCODE_eQTLs | *ACP6* | 1.50e-07 |
| rs1891505 | 1 | 147208624 | 0.26 | *GJA5* | 5.417e-09 | eQTLGen_cis_eQTLs | *ACP6* | 1.24e-07 |
| rs1539418 | 1 | 147210110 | 0.26 | *GJA5* | 5.488e-09 | eQTLGen_cis_eQTLs | *ACP6* | 1.97e-07 |
| rs7514182 | 1 | 147207229 | 0.27 | *GJA5* | 6.617e-09 | PsychENCODE_eQTLs | *ACP6* | 3.32e-07 |
| rs11590206 | 1 | 147199309 | 0.28 | *RP11-533N14.3* | 7.2e-09 | PsychENCODE_eQTLs | *ACP6* | 1.18e-10 |
| rs11590206 | 1 | 147199309 | 0.28 | *RP11-533N14.3* | 7.2e-09 | eQTLGen_cis_eQTLs | *ACP6* | 5.33e-11 |
| rs11576092 | 1 | 147198761 | 0.28 | *RP11-533N14.3* | 8.706e-09 | eQTLGen_cis_eQTLs | *ACP6* | 5.26e-11 |
| rs10900396 | 1 | 147242596 | 0.29 | *GJA5* | 1.492e-08 | PsychENCODE_eQTLs | *ACP6* | 02.7e-4 |
| rs1857207 | 1 | 147251276 | 0.18 | *RP11-433J22.3* | 1.716e-08 | PsychENCODE_eQTLs | *ACP6* | 3.00e-4 |
| rs1857207 | 1 | 147251276 | 0.18 | *RP11-433J22.3* | 1.716e-08 | eQTLGen_cis_eQTLs | *ACP6* | 1.02e-11 |
| rs8179281 | 1 | 147308112 | 0.18 | *RP11-314N2.2* | 2.015e-08 | eQTLGen_cis_eQTLs | *ACP6* | 3.08e-12 |
| rs10900397 | 1 | 147243153 | 0.22 | *GJA5* | 2.078e-08 | PsychENCODE_eQTLs | *ACP6* | 3.99e-06 |
| rs10900397 | 1 | 147243153 | 0.22 | *GJA5* | 2.078e-08 | eQTLGen_cis_eQTLs | *ACP6* | 2.30e-13 |
| rs11240122 | 1 | 147235547 | 0.27 | *GJA5* | 3.934e-08 | PsychENCODE_eQTLs | *ACP6* | 6.60e-4 |
| rs12409049 | 1 | 147236467 | 0.27 | *GJA5* | 3.934e-08 | PsychENCODE_eQTLs | *ACP6* | 6.92e-4 |
| rs12023381 | 1 | 147265367 | 0.20 | *RP11-433J22.3* | 6.438e-08 | eQTLGen_cis_eQTLs | *ACP6* | 1.64e-13 |
| rs10494256 | 1 | 147296184 | 0.18 | *RP11-314N2.2* | 1.143e-07 | eQTLGen_cis_eQTLs | *ACP6* | 2.52e-12 |
| rs12034574 | 1 | 147292366 | 0.18 | *RP11-314N2.2* | 1.208e-07 | eQTLGen_cis_eQTLs | *ACP6* | 6.71e-12 |
| rs12035481 | 1 | 147301477 | 0.18 | *RP11-314N2.2* | 1.26e-07 | PsychENCODE_eQTLs | *ACP6* | 1.44e-05 |
| rs12035481 | 1 | 147301477 | 0.18 | *RP11-314N2.2* | 1.26e-07 | eQTLGen_cis_eQTLs | *ACP6* | 7.10e-12 |
| rs72702336 | 1 | 147290056 | 0.18 | *RP11-314N2.2* | 1.314e-07 | PsychENCODE_eQTLs | *ACP6* | 2.08e-05 |
| rs72702336 | 1 | 147290056 | 0.18 | *RP11-314N2.2* | 1.314e-07 | eQTLGen_cis_eQTLs | *ACP6* | 1.30e-12 |
| rs59470904 | 1 | 147288501 | 0.18 | *RP11-314N2.2* | 1.439e-07 | PsychENCODE_eQTLs | *ACP6* | 4.22e-06 |
| rs59470904 | 1 | 147288501 | 0.18 | *RP11-314N2.2* | 1.439e-07 | eQTLGen_cis_eQTLs | *ACP6* | 9.58e-13 |
| rs1857208 | 1 | 147291987 | 0.18 | *RP11-314N2.2* | 1.484e-07 | eQTLGen_cis_eQTLs | *ACP6* | 5.72e-13 |
| rs72702335 | 1 | 147289061 | 0.18 | *RP11-314N2.2* | 2.108e-07 | PsychENCODE_eQTLs | *ACP6* | 4.10e-06 |
| rs72702335 | 1 | 147289061 | 0.18 | *RP11-314N2.2* | 2.108e-07 | eQTLGen_cis_eQTLs | *ACP6* | 1.32e-12 |
| rs6667651 | 1 | 147170417 | 0.26 | *RP11-533N14.3* | 2.436e-07 | PsychENCODE_eQTLs | *ACP6* | 3.344e-16 |
| rs6667651 | 1 | 147170417 | 0.26 | *RP11-533N14.3* | 2.436e-07 | eQTLGen_cis_eQTLs | *ACP6* | 7.36e-21 |
| rs6701830 | 1 | 147170542 | 0.26 | *RP11-533N14.3* | 2.476e-07 | PsychENCODE_eQTLs | *ACP6* | 3.38e-16 |
| rs6701830 | 1 | 147170542 | 0.26 | *RP11-533N14.3* | 2.476e-07 | eQTLGen_cis_eQTLs | *ACP6* | 1.98e-20 |
| rs34739004 | 1 | 147192029 | 0.24 | *RP11-533N14.3* | 2.518e-07 | PsychENCODE_eQTLs | *ACP6* | 1.26e-16 |
| rs34739004 | 1 | 147192029 | 0.24 | *RP11-533N14.3* | 2.518e-07 | eQTLGen_cis_eQTLs | *ACP6* | 2.18e-29 |
| rs34739004 | 1 | 147192029 | 0.24 | *RP11-533N14.3* | 2.518e-07 | Brain_Caudate_basal_ganglia | *ACP6* | 3.69e-05 |
| rs7536576 | 1 | 147310661 | 0.19 | *RP11-314N2.2* | 2.694e-07 | PsychENCODE_eQTLs | *ACP6* | 6.75e-05 |
| rs7536576 | 1 | 147310661 | 0.19 | *RP11-314N2.2* | 2.694e-07 | eQTLGen_cis_eQTLs | *ACP6* | 2.69e-12 |
| rs112858789 | 1 | 147300422 | 0.18 | *RP11-314N2.2* | 3.816e-07 | PsychENCODE_eQTLs | *ACP6* | 2.06e-06 |
| rs112858789 | 1 | 147300422 | 0.18 | *RP11-314N2.2* | 3.816e-07 | eQTLGen_cis_eQTLs | *ACP6* | 1.30e-15 |

| SNP | CHR | POS | MAF | nearestGene | gwasP | tissue | symbol | eqtlP |
|---|---|---|---|---|---|---|---|---|
| rs11582163 | 1 | 147167854 | 0.25 | *RP11-533N14.3* | 4.624e-07 | eQTLGen_cis_eQTLs | *ACP6* | 2.94e-33 |
| rs11583679 | 1 | 147167893 | 0.25 | *RP11-533N14.3* | 4.624e-07 | PsychENCODE_eQTLs | *ACP6* | 5.31e-17 |
| rs11583679 | 1 | 147167893 | 0.25 | *RP11-533N14.3* | 4.624e-07 | eQTLGen_cis_eQTLs | *ACP6* | 3.22e-33 |
| rs10900399 | 1 | 147283261 | 0.20 | *RP11-433J22.3* | 4.699e-07 | eQTLGen_cis_eQTLs | *ACP6* | 4.57e-11 |
| rs11586163 | 1 | 147165272 | 0.26 | *RN7SL261P* | 4.983e-07 | eQTLGen_cis_eQTLs | *ACP6* | 1.61e-20 |
| rs4950346 | 1 | 147282446 | 0.31 | *RP11-433J22.3* | 5.318e-07 | PsychENCODE_eQTLs | *ACP6* | 07.29e-4 |
| rs4950346 | 1 | 147282446 | 0.31 | *RP11-433J22.3* | 5.318e-07 | PsychENCODE_eQTLs | *LINC01138* | 5.07e-4 |
| rs11585427 | 1 | 147168846 | 0.25 | *RP11-533N14.3* | 7.523e-07 | eQTLGen_cis_eQTLs | *ACP6* | 6.44e-33 |
| rs12747247 | 1 | 147166010 | 0.25 | *RP11-533N14.3* | 8.056e-07 | PsychENCODE_eQTLs | *ACP6* | 7.76e-17 |
| rs12747247 | 1 | 147166010 | 0.25 | *RP11-533N14.3* | 8.056e-07 | eQTLGen_cis_eQTLs | *ACP6* | 5.25e-33 |
| rs1001193 | 1 | 147166377 | 0.25 | *RP11-533N14.3* | 8.056e-07 | eQTLGen_cis_eQTLs | *ACP6* | 7.68e-33 |
| rs11586154 | 1 | 147165238 | 0.25 | *RN7SL261P* | 1.071e-06 | PsychENCODE_eQTLs | *ACP6* | 2.165e-17 |
| rs11586154 | 1 | 147165238 | 0.25 | *RN7SL261P* | 1.071e-06 | eQTLGen_cis_eQTLs | *ACP6* | 1.34e-34 |
| rs10900400 | 1 | 147314153 | 0.17 | *RP11-314N2.2* | 2.888e-06 | eQTLGen_cis_eQTLs | *ACP6* | 2.12e-17 |
| rs791297 | 1 | 147285656 | 0.44 | *RP11-433J22.3* | 1.002e-05 | PsychENCODE_eQTLs | *LINC01138* | 6.69e-4 |
| rs4950336 | 1 | 147174692 | 0.33 | *RP11-533N14.3* | 0.0007038 | eQTLGen_cis_eQTLs | *BCL9* | 2.38e-06 |
| rs4950336 | 1 | 147174692 | 0.33 | *RP11-533N14.3* | 0.0007038 | eQTLGen_cis_eQTLs | *ACP6* | 4.37e-06 |
| rs11589555 | 1 | 147183232 | 0.34 | *RP11-533N14.3* | 0.0008416 | PsychENCODE_eQTLs | *ACP6* | 1.87e-09 |
| rs1342710 | 1 | 147173505 | 0.33 | *RP11-533N14.3* | 0.0008491 | eQTLGen_cis_eQTLs | *BCL9* | 2.27e-06 |
| rs1342710 | 1 | 147173505 | 0.33 | *RP11-533N14.3* | 0.0008491 | eQTLGen_cis_eQTLs | *ACP6* | 1.71e-05 |
| rs12760985 | 1 | 147186616 | 0.34 | *RP11-533N14.3* | 0.0009745 | PsychENCODE_eQTLs | *ACP6* | 1.06e-09 |
| rs4405176 | 1 | 147181058 | 0.33 | *RP11-533N14.3* | 0.0009836 | PsychENCODE_eQTLs | *ACP6* | 2.05e-09 |
| rs4405176 | 1 | 147181058 | 0.33 | *RP11-533N14.3* | 0.0009836 | eQTLGen_cis_eQTLs | *BCL9* | 3.39e-06 |
| rs4405176 | 1 | 147181058 | 0.33 | *RP11-533N14.3* | 0.0009836 | eQTLGen_cis_eQTLs | *ACP6* | 7.158e-06 |
| rs11587198 | 1 | 147173713 | 0.33 | *RP11-533N14.3* | 0.001047 | PsychENCODE_eQTLs | *ACP6* | 1.17e-09 |
| rs11587198 | 1 | 147173713 | 0.33 | *RP11-533N14.3* | 0.001047 | eQTLGen_cis_eQTLs | *BCL9* | 3.87e-06 |
| rs11587198 | 1 | 147173713 | 0.33 | *RP11-533N14.3* | 0.001047 | eQTLGen_cis_eQTLs | *ACP6* | 1.08e-05 |
| rs11590104 | 1 | 147184498 | 0.34 | *RP11-533N14.3* | 0.001081 | PsychENCODE_eQTLs | *ACP6* | 1.078e-09 |
| rs12755420 | 1 | 147185553 | 0.34 | *RP11-533N14.3* | 0.00132 | PsychENCODE_eQTLs | *ACP6* | 1.11e-09 |

#### eFigure 1. Quality Control flowchart

| Level | QC step | TPD | OPDC | PPMI | PDSTAT | DIGPD | PDBP |
|---|---|---|---|---|---|---|---|
| Clinical QC | N | 2000 | 1082 | 415 | 174 | 427 | 873 |
| Clinical QC | Longitudinal Clinical & genotype data available & no duplicates | 1824 | 872 | 413 | 128 | 423 | 360 |
| Sample level QC | Sex pass | 1819 | 870 | 412 | 128 | 423 | 360 |
| Sample level QC | Sample missing rate >2% & heterozygosity rate > ±3SD from mean | 1780 | 845 | 404 | 126 | 382 | 360 |
| Sample level QC | PIHAT > 0.0875 | 1767 | 831 | 396 | 124 | 382 | 360 |
| Sample level QC | Ancestry | 1717 | 803 | 287 | 124 | 376 | 360 |
| Sample level QC | Further sample missing rate and homozygosity rate filtering | 1700 | 797 | 287 | 124 | 374 | 360 |
| Sample level QC | Further PIHAT filtering | 1699 | 797 | 287 | 124 | 374 | 360 |
| Variant level QC | N | 266152 | 557018 | 61715058 | 476011 | 1778953 | 158715874 |
| Variant level QC | Genotyping missing rate > 5% | 266152 | 513158 | 58129433 | 431276 | 1762893 | 158546878 |
| Variant level QC | MAF < 0.01 | 265622 | 257801 | 11370163 | 431276 | 835782 | 11723465 |
| Variant level QC | mishap | 265622 | 257193 | 11335953 | 223371 | 835416 | 11723465 |
| Variant level QC | HWE < 1e-10 | 265622 | 257159 | 11182129 | 223371 | 835286 | 11203502 |
| Pre-imputation | Genotype rate | 0.999 | 0.999 | 0.999 | 0.999 | 0.999 | 0.999 |
| Post-imputation QC | R^2 > 0.8 | 6864740 | 6727127 | 11182129 | 6429450 | 11440561 | 11203502 |
| Post-imputation QC | Genotyping missing rate >0.05 | 6754740 | 6219170 | 11182129 | 5036951 | 11440561 | 11203502 |
| Post-imputation QC | MAF > 0.01 | 6754740 | 6219170 | 11182129 | 5036951 | 7335865 | 11203502 |
| TOTAL | Total SNPs post-imputation | 6754740 | 6219170 | 11182129 | 5036951 | 7335865 | 11203502 |

Each row shows a different QC step at different levels ( Clinical QC, Sample level QC, Variant level QC, Post-Imputation QC) across each cohort displayed as a column. In addition, a metric of the genotyping rate prior imputation is shown in Pre-imputation. The resulting number of SNPs available for the study in each cohort is shown in TOTAL.

**eFigure 2. Equations predicting the impact of levodopa dosage in MDS-UPDRS part III.**

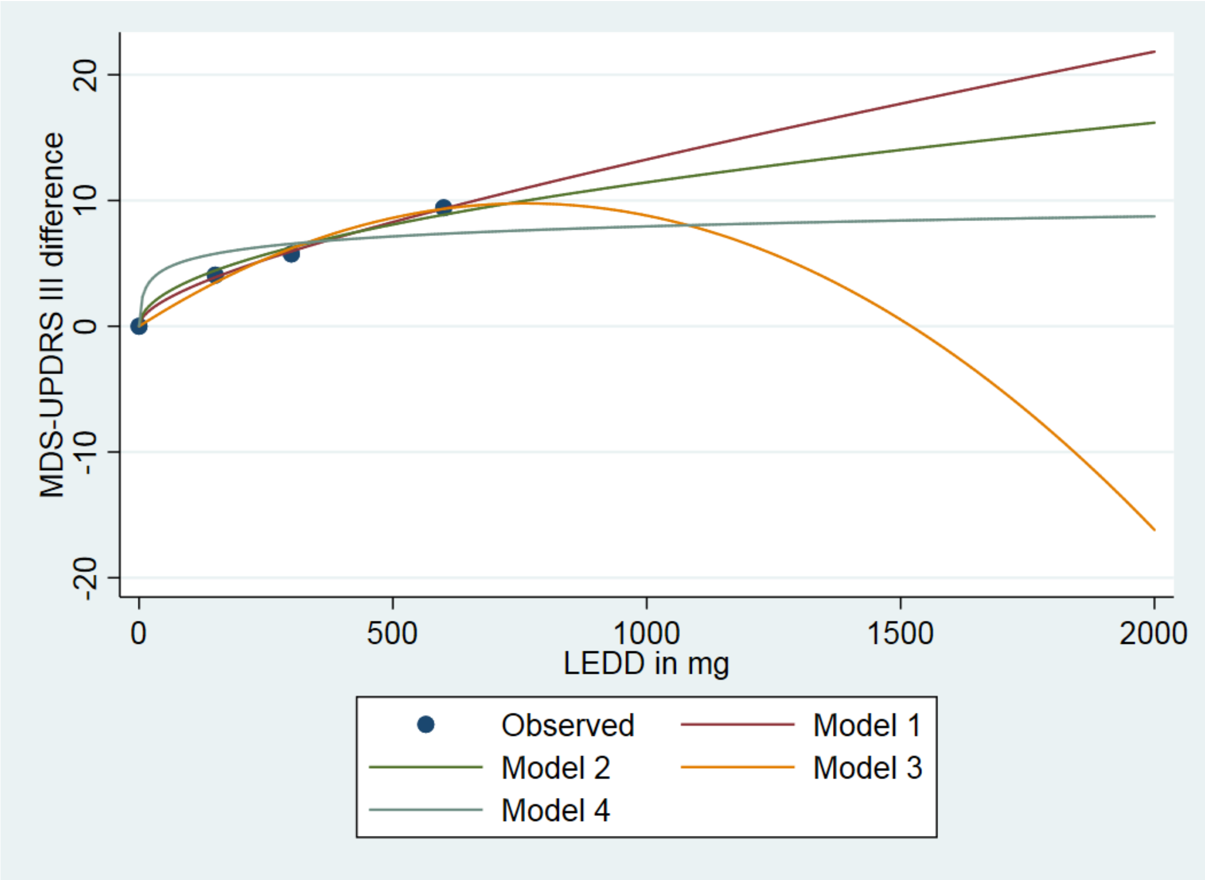


Model 1 = Model with linear and square root term. Model 2 = Model with square root term only, Model 3 = Model with linear and square term, Model 4 = Model with a log term (where 1 is added to the x-axis since log is undefined at a value of 0). All the curves were derived so that they have no constant term which ensures that at a LEDD of zero that curve passes through zero.All the curves had an R-square > 0.99 except model 4. We see how model 1 and model 2 extrapolate the data best past an LEDD of 600mg. We concluded model 2 is what we would expect to happen at higher LEDD increasing slightly slower.

### eFigure 3. LMMs Assumptions checking

a)

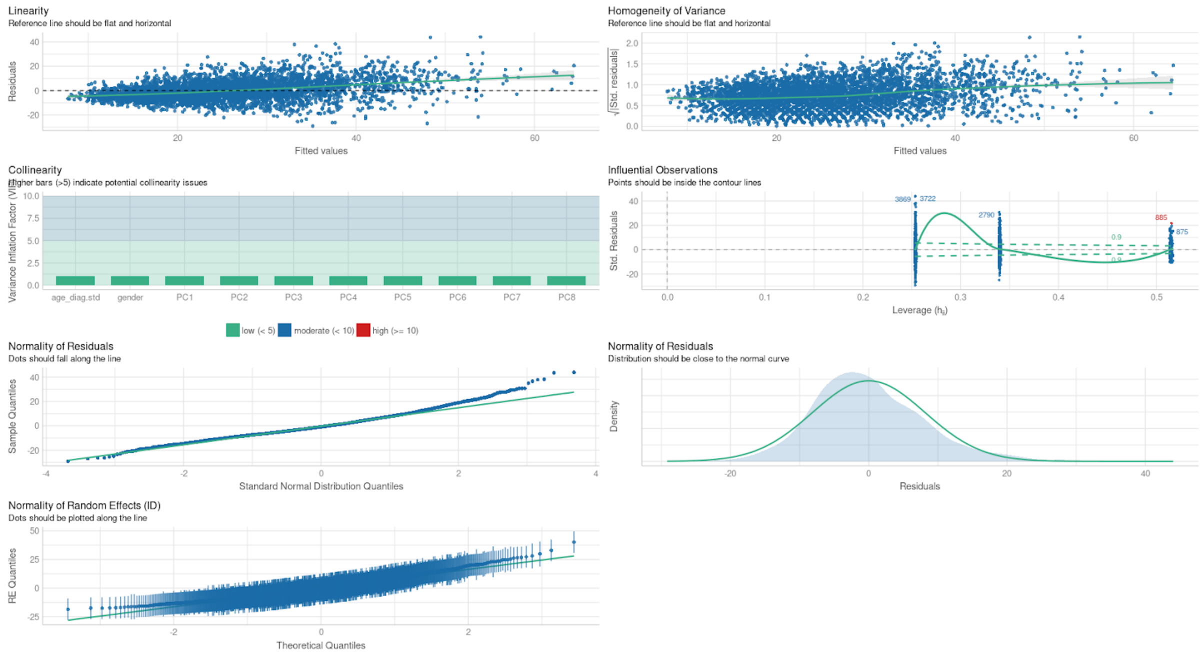


b)

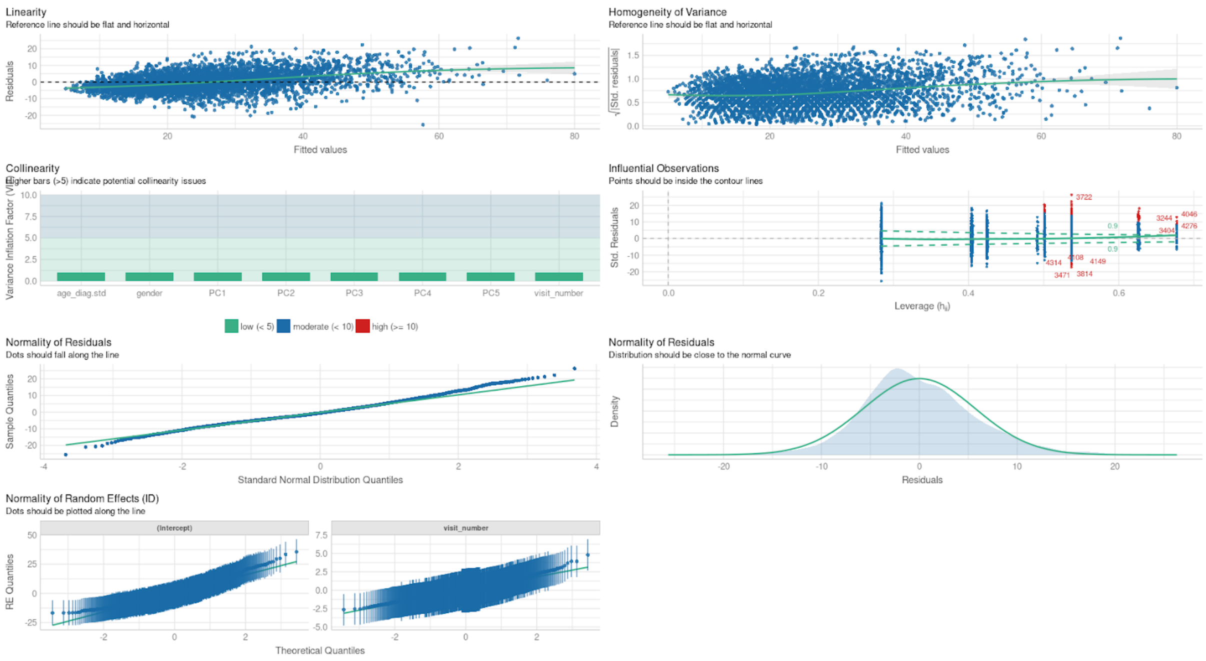


a-b) Checking assumptions necessary for models A and B respectively. **Each panel tests one of the following assumptions:** Linearity, homogeneity of variance assumption, collinearity, Influential observations, normality of residuals, and Normality of random effects assumption.

#### eFigure 4. SCEBE validation in OPDC and DIGPD cohorts.

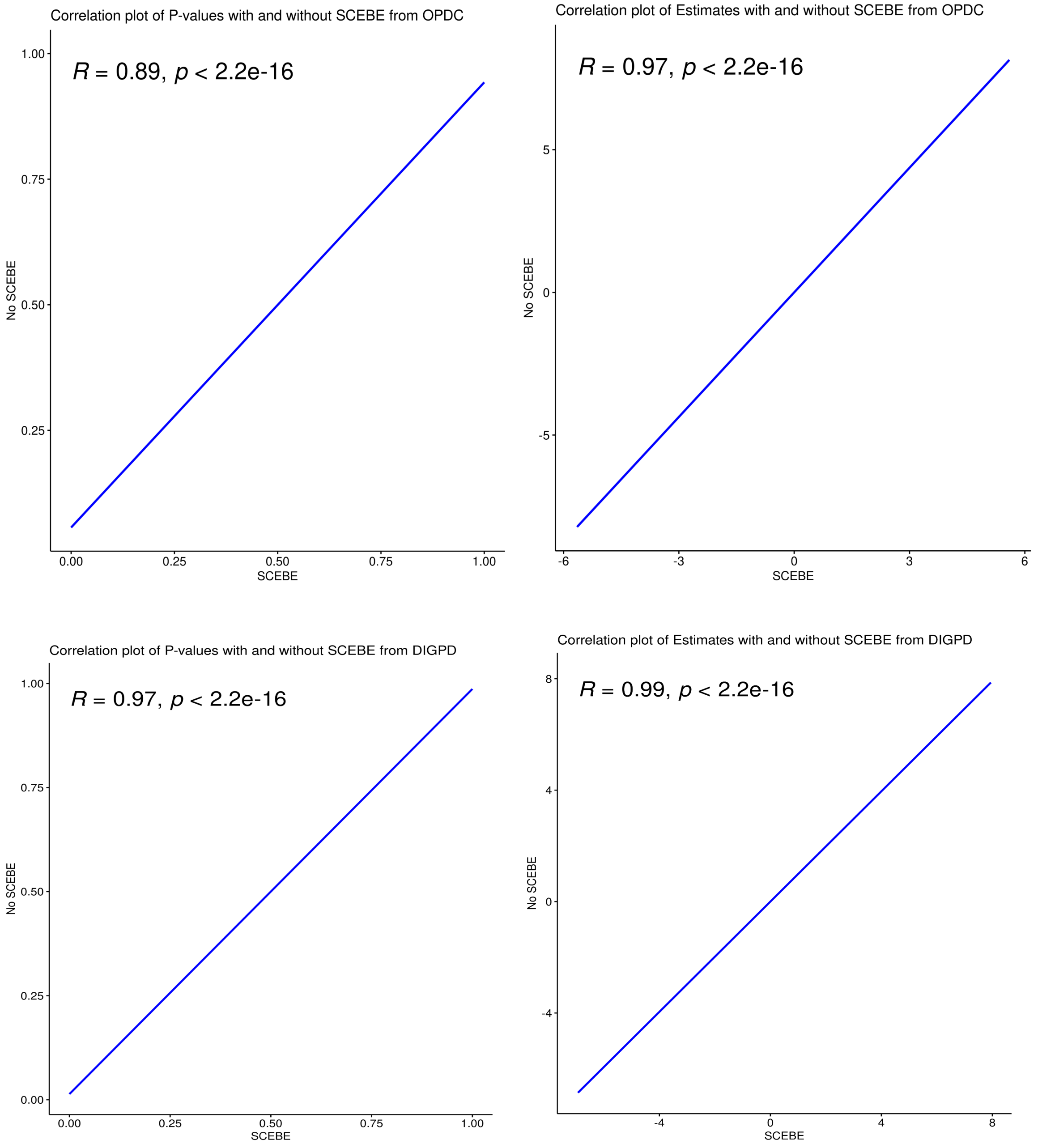


Correlation plots between P-values and Coefficients derived with SCEBE approach (X-axis) and with lmeRTest using the Satterwhite approach to derive P-values (Y-axis). The two top figures are the correlation plots of models fitted with OPDC data. The two bottom figures are the correlation plots of models fitted using DIGPD data. We used MDS-UPDRS III total as the outcome of the model. Each plot shows the correlation value ($R^2$), and the significance of the correlation (*P*).

#### eFigure 5. Power to detect genetic associations in LMMs.

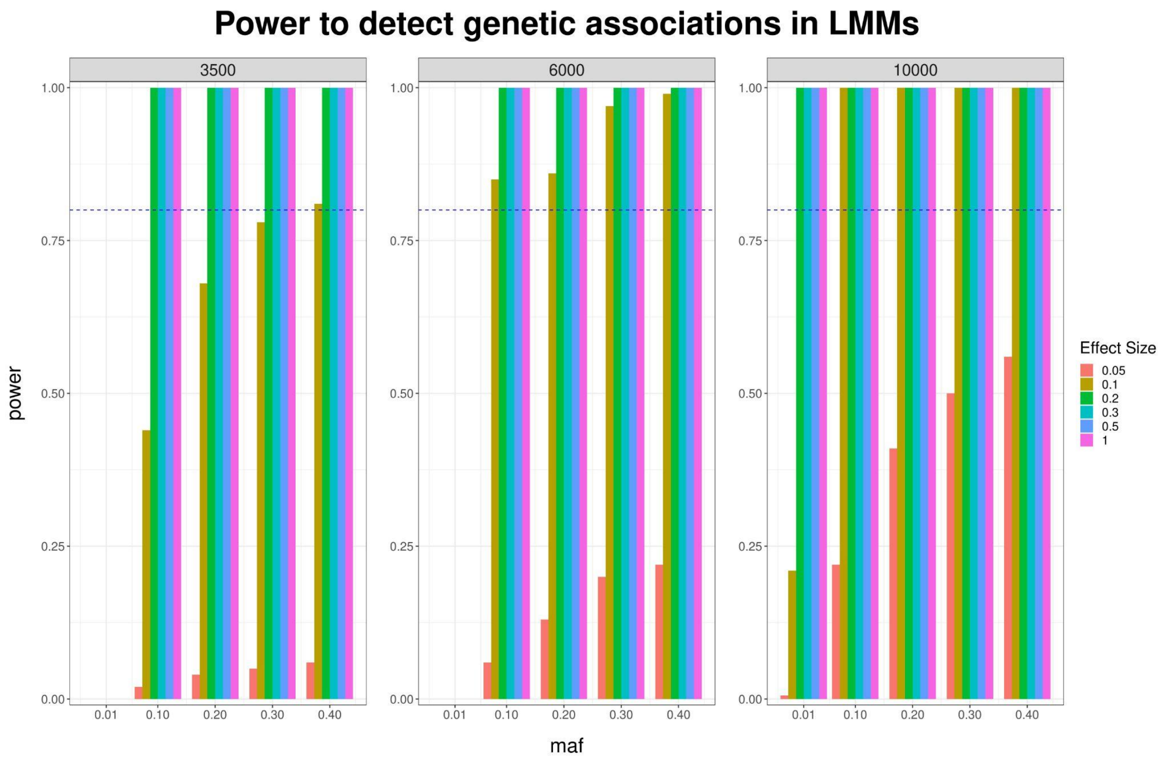


The Y axis shows the power (0 to 1). The X axis shows the MAF of the SNP tested 10000 times. The header of each plot represents the sample size. Different colours represent the simulated effect size.

### eFigure 6. Fine Mapping and regional plots of MAD1L1 and LINC00511 loci.

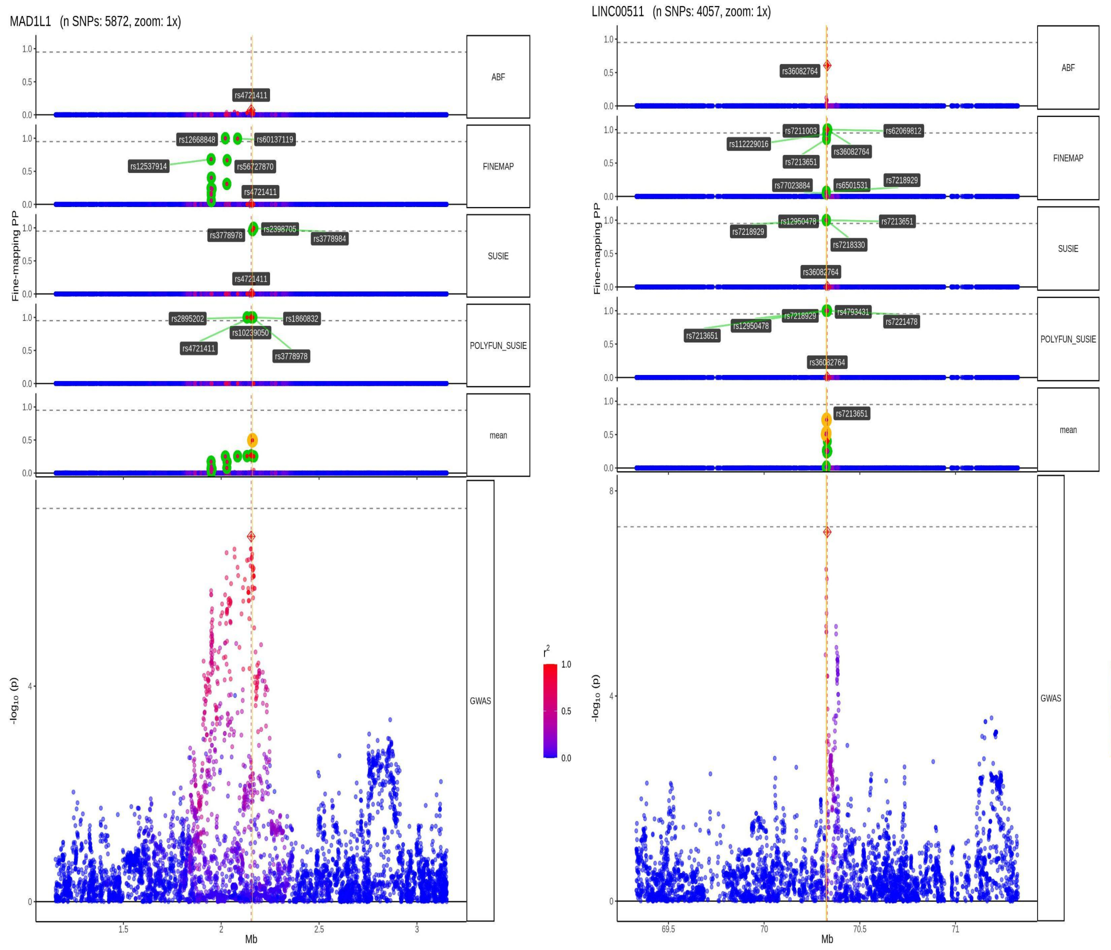


Fine Mapping output and regional plots of *MAD1L1* (left), and *LINC00511* (right) loci. The first five rows show the cross tools fine-mapping output ( ABF, FINEMAP, SUSIE, and POLYFUN-SUSIE) and the Consensus variants (support from at least two fine-mapping tools). We can see the per SNP ( x-axis) Posterior Probability from different tools ( y-axis). At the bottom we see the log(10) *P* ( y axis) of SNPs on each locus.

#### eFigure 7. MAD1L1 Regional plots from axial motor GWAS and PsychEncode cis-eQTL

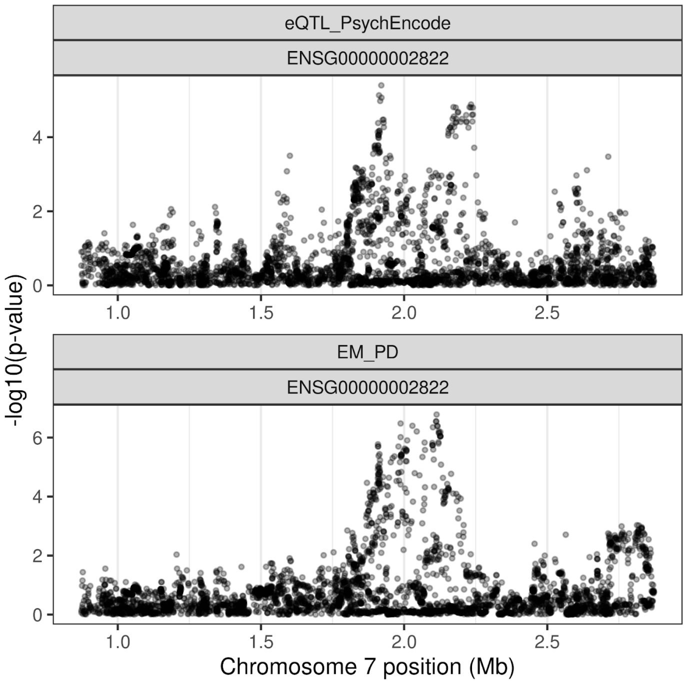


#### eFigure 8. Manhattan plot for model B GWAS meta-analysis using HY as the outcome.

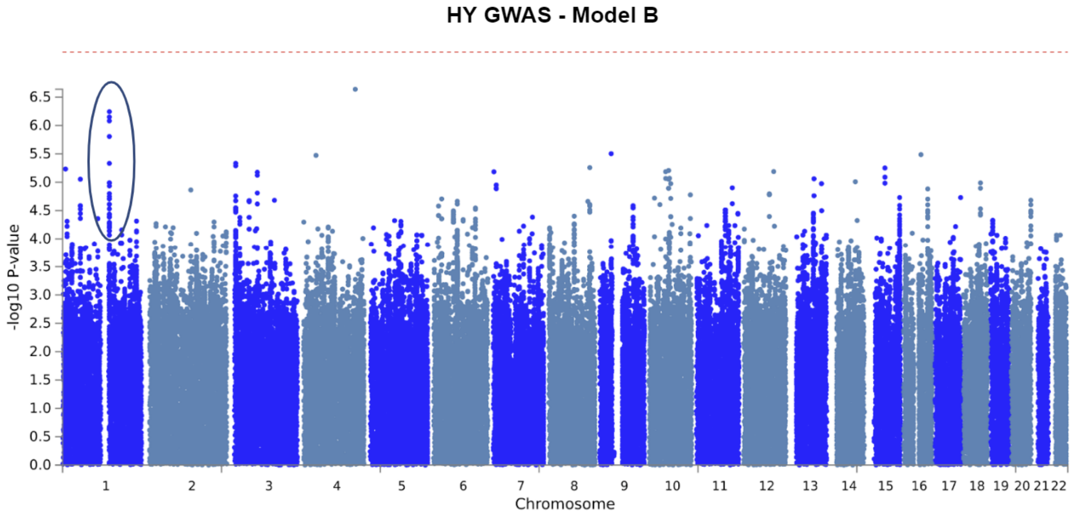


GWAS meta-analysis manhattan plot  for HY outcome under model B described in methods.
